# Cost-Effective Threshold Price for Alternative Infant and Neonatal Rotavirus Vaccines: A Dual-Country Evaluation

**DOI:** 10.64898/2026.05.12.26353029

**Authors:** Xiao Li, Ernest O. Asare, Jiye Kwon, Catherine G. C. Wenger, George E. Armah, Nigel A. Cunliffe, Khuzwayo C. Jere, Joke Bilcke, Philippe Beutels, Virginia E. Pitzer

## Abstract

Suboptimal rotavirus vaccine effectiveness in low- and middle-income countries (LMICs) highlights the need for next-generation vaccines, such as the neonatal RV3-BB vaccine. However, there is uncertainty in the duration of protection and future price of vaccines in development. We aim to identify the conditions under which switching to RV3-BB is optimal in Malawi and Ghana, where the current immunization programs use 2-dose Rotarix and 3-dose Rotavac schedules, respectively. A full incremental cost-effectiveness analysis was performed using a validated transmission model calibrated to country-specific rotavirus data. Over 2025–2034, introducing RV3-BB resulted in the largest rotavirus-related burden reduction compared with the current country-specific programs. At moderate willingness-to-pay (∼0.5-time Gross Domestic Product per capita), RV3-BB was preferred over Rotavac if price per dose was <$1.2 in Malawi and <$2.5 in Ghana, and/or if the average duration of protection exceeded 40 weeks in Malawi. The RV3-BB vaccine is likely to be cost-effective in Malawi and Ghana, as well as other LMICs, based on expected pricing and duration of protection.

## 1 Introduction

Despite major reductions in under-five mortality rates over recent decades ^1^, diarrheal disease remains the leading cause of childhood deaths in low- and middle-income countries (LMICs) ^2^. The introduction of rotavirus vaccines has markedly reduced the incidence of rotavirus gastroenteritis (RVGE) worldwide, with substantial impacts on preventing severe outcomes, including premature deaths ^3,4^. Nevertheless, RVGE remains a leading cause of diarrheal mortality and continues to impose a substantial public health burden among children, particularly in settings where health care resources and access to timely care are limited ^5^.

In 2009, the WHO recommended routine rotavirus vaccination of infants worldwide, and Gavi began supporting introduction through the global rotavirus vaccination program in 2012. Rotavirus vaccination has been widely implemented within national immunization programs (NIPs) across 134 countries, including 45 Gavi-eligible countries by 2025 ^6^.

Four oral rotavirus vaccines are pre-qualified by the World Health Organization (WHO) for global use ^7^. In 2009, Rotarix^®^ (GSK) vaccine, using a 2-dose schedule (at 6 and 10 or 10 and 14 weeks of age), and RotaTeq^®^ (Merck) vaccine, using a 3-dose schedule (at 6, 10, and 14 weeks) were recommended. In January 2018, Rotavac^®^ (Bharat Biotech) vaccine, using a 3-dose schedule (at 6, 10, and 14 weeks), received WHO prequalification ^7,8^. In September 2018, another 3-dose vaccine, Rotasiil^®^ (Serum Institute of India), also received WHO prequalification with the same 3-dose schedule^7^. Both Rotavac^®^ and Rotasiil^®^ offered a markedly lower Gavi-negotiated price per dose than Rotarix^®^ and RotaTeq^®^, and RotaTeq has been withdrawn from the Gavi market since 2019.

Rotavirus vaccines have shown lower efficacy in LMICs than in high-income countries (HICs), which may be partly related to the earlier occurrence of severe RVGE in these settings ^5,9,10^. Other factors that have been suggested to contribute include differences in enteric pathogen exposure and gut microbiome composition, and potential interactions with co-administered vaccines such as oral polio vaccine^11,12^. likely associated with a higher occurrence of in LMICs. As a result, new vaccines and alternative dosing schedules are being investigated to enhance protection among vulnerable children. A promising new neonatal vaccine candidate, RV3-BB, is currently undergoing clinical trials ^13,14^. Delivered in an advanced 3-dose schedule at birth, 6, and 10 weeks of age, it can offer protection earlier in life and may therefore further reduce the high rotavirus disease burden occurring in infancy in many LMICs. Nonetheless, uncertainty remains regarding the effectiveness and protective duration of the neonatal schedule. Moreover, alternative strategies are being explored, for instance, switching from a 2-dose Rotarix schedule to a 3-dose Rotavac schedule, or adding a third dose of Rotarix to improve immunogenicity compared with the standard 2-dose Rotarix schedule^15–17^.

Economic evaluation has been widely used to inform decision-making, particularly when considering the introduction of a new vaccine or a switch to an alternative product or dosing schedule ^18,19^. Assessing the cost-effectiveness of new immunization strategies relative to the current program is critical for understanding their value and guiding policy choices ^18^. Moreover, it is also essential to account for uncertainty around the cost-effective strategy and the potential opportunity costs if the wrong choice is made^20–23^.

The objectives of this study are (1) to identify the conditions under which switching to the neonatal vaccine or switching to a different dosing schedule may be the optimal rotavirus vaccination strategy from the government and societal perspectives, and (2) to identify the key drivers of decision uncertainty. We applied these analyses to two countries for illustrative purposes. Malawi, a low-income Southeastern African country, introduced the 2-dose Rotarix vaccine in 2012 and continues to use it in its NIP. Ghana, a lower-middle-income West African country, introduced the 2-dose Rotarix schedule in 2012 but switched to the 3-dose Rotavac schedule in 2020.

## 2 Results

### 2.1 Vaccination Strategies and Comparative Analysis

We considered rotavirus vaccination strategies over a 10-year time horizon from 2025 to 2034 (Table 1). In Malawi, where the 2-dose Rotarix schedule serves as the baseline, we compared four strategies: (1) suspension of vaccination in 2025, (2) adding a third Rotarix dose at 14 weeks of age, (3) switching to the 3-dose Rotavac schedule, and (4) adopting the neonatal RV3-BB schedule administered at 1, 6, and 10 weeks.

**Table 1:** Country-specific vaccination strategies evaluated.

| Vaccination program | Malawi | Ghana |
| --- | --- | --- |
| Baseline (current national immunisation program) | Rotarix 2-dose schedule: 6/10 weeks of age | Rotavac 3-dose schedule: 6/10/14 weeks of age |
| Suspension of rotavirus vaccination program | Suspending vaccination | Suspending vaccination |
| Rotarix 3-dose schedule | Rotarix: 6/10/14 weeks | Not relevant |
| Rotavac 3-dose schedule | Rotavac: 6/10/14 weeks | Baseline |
| Neonatal RV3-BB vaccine<br>3-dose schedule | RV3BB: 1/6/10 weeks | RV3-BB: 1/6/10 weeks |

In Ghana, where the 3-dose Rotavac schedule has been implemented since 2020, a switch back to a Rotarix program (2-dose or 3-dose) seems unlikely ^24^. We therefore compared only two strategies to the baseline strategy: suspension of vaccination and switching to the neonatal RV3-BB strategy.

### 2.2 Predicted rotavirus disease and economic burden over 10 years by vaccination strategy

Introducing RV3-BB produced the largest reductions in rotavirus disease burden in both countries (Figure S.1). In Malawi, RV3-BB reduced moderate-to-severe and non-severe cases by 18% and 11%, respectively, outperforming alternative strategies such as adding a third Rotarix dose or switching to Rotavac. In Ghana, switching from the current Rotavac schedule to RV3-BB reduced moderate-to-severe cases by 25% and non-severe cases by 15%. In contrast, suspending rotavirus vaccination substantially increased disease burden in both settings, with large increases in RVGE cases, especially moderate-to-severe cases.

Consequently, introducing RV3-BB in Malawi and Ghana reduced RVGE-associated deaths and hospitalizations by approximately 18–25% and outpatient and non– medically attended (non-MA) cases by 11–16%, whereas suspending vaccination increased disease burden by 39–42% for severe outcomes and 14–41% for non-hospitalized cases (Table S.5).

When excluding vaccine-related costs, the discounted RVGE-related costs and disability-adjusted life-years (DALYs) are shown in Figure S.2 by vaccination strategy. Compared with the current NIP, RV3-BB consistently yielded the lowest discounted costs and DALYs, while suspension of vaccination resulted in markedly large increases in costs and DALYs.

The discounted incremental direct and total costs and DALYs averted by strategy and country are presented in Table S.6 at vaccine prices of $1.79\comma \; $0.70, and $0.70 per dose for Rotarix\comma \; Rotavac\comma \; and RV3-BB\comma \; respectively. Incremental analyses showed that on average\comma \; introducing RV3-BB was associated with additional health gains of 44 thousand DALYs averted\comma \; and savings of $6 million direct cost from the government perspective, and $8 million total costs from a societal perspective when compared with the current NIP in Malawi. In Ghana\comma \; RV3-BB averted 87 thousand DALYs and led to mean savings of $1 million direct costs from the government perspective, and $5 million total costs from a societal perspective. In contrast\comma \; suspending rotavirus vaccination consistently reduced direct costs by $22 million in Malawi and $64 million in Ghana due to reduced spending on vaccination. However\comma \; it also resulted in substantial increases in disease burden\comma \; with 95 and 150 thousand additional DALYs. In Malawi\comma \; alternative strategies\comma \; such as adding a third Rotarix dose\comma \; increased direct costs by approximately $11 million while averting 39 thousand DALYs, whereas switching to Rotavac yielded moderate direct cost savings ($2 million) and health gains (33 thousand DALYs).

### 2.3 Full incremental cost-effectiveness analysis

The cost-effectiveness planes for the full incremental analysis are presented in Figures S.3 and S.4 from both government and societal perspectives. For both countries at a vaccine price of $0.70\comma \; RV3-BB dominated the other options\comma \; with expected incremental cost-effectiveness ratios \lpar ICERs\rpar of $117 (Malawi) and $266 \lpar Ghana\rpar per DALY averted\comma \; compared to suspending rotavirus vaccination. For willingness-to-pay \lpar WTP\rpar values lower than these ICERs\comma \; suspending vaccination was preferred. ICERs from the societal perspective were lower than those from the government perspective \lpar $74 in Malawi and $192 in Ghana per DALY averted) when compared with suspension of rotavirus vaccination.

For both Malawi and Ghana, results from the cost-effectiveness acceptability curves and frontiers (CEAC and CEAF, respectively), expected net loss (ENL), and expected value of perfect information (EVPI) analyses are presented in Figure 1 and Figure S.5 from the government and societal perspectives, respectively. Decision uncertainty is large around WTP values equal to these ICERs, and ENL was estimated around $10 million for both countries. In Ghana\comma \; from WTP values above $600 per DALY averted, RV3-BB is preferred with close to absolute certainty (probability cost-effective >99% and ENL $10\comma \; 000\rpar . In Malawi\comma \; Rotavac shows 30&percnt\semicolon \; probability of being preferred at WTP values \gt $250 per DALY averted. The 2-dose (current NIP) and 3-dose Rotarix schedules consistently showed lower probabilities of being cost-effective and higher expected net losses than Rotavac and RV3-BB, largely due to their higher price per dose; their probability of being cost-effective and uncertainty (ENL) increased gradually as WTP values rose. This indicated the importance of the RV3-BB price sensitivity analyses.

**Figure 1:**
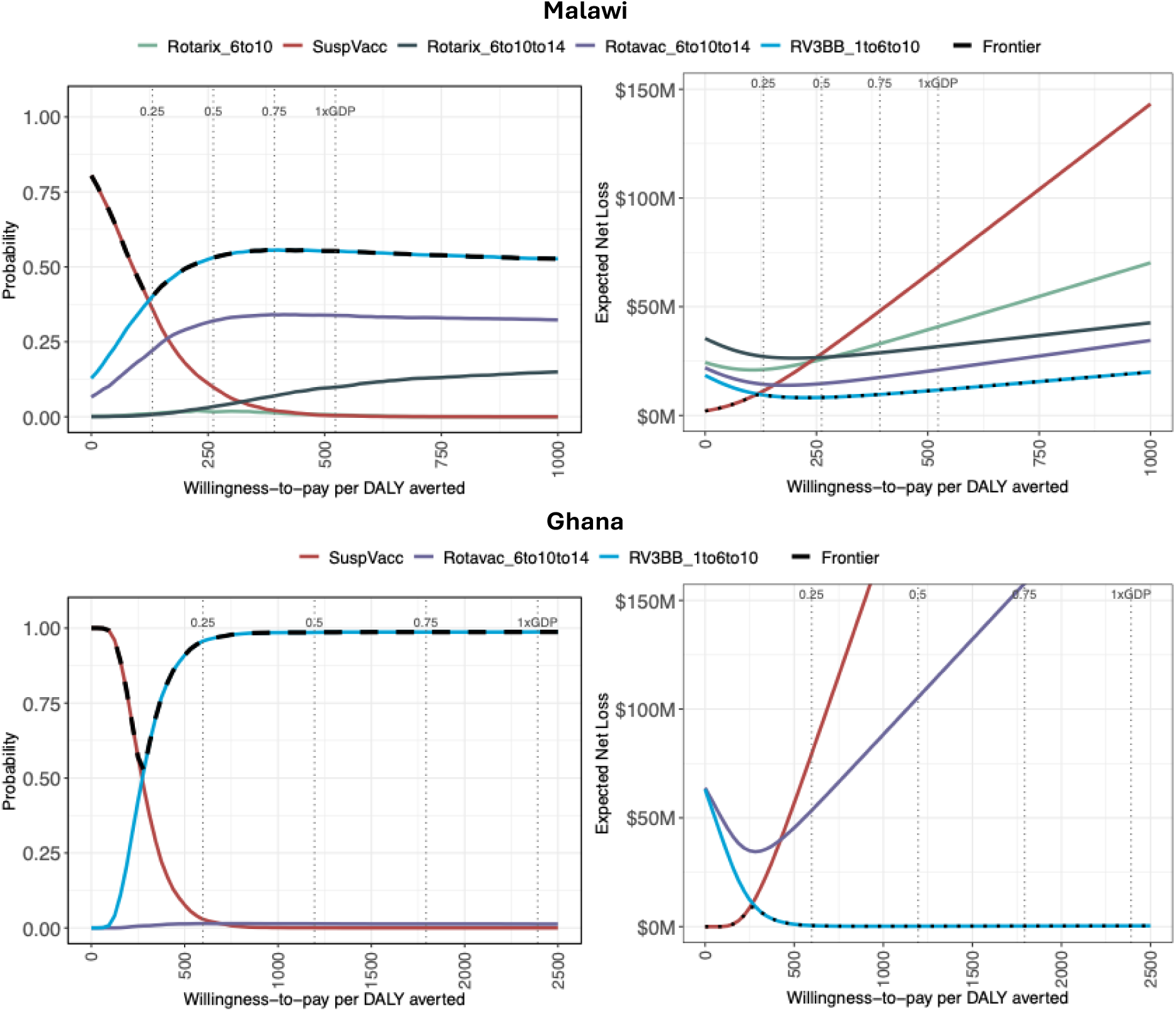
**Cost-effectiveness acceptability curve, cost-effectiveness acceptability frontier, expected net losses and expected value of information for Malawi (top panel) and Ghana (bottom panel)**. All the relevant strategies were compared with the current national immunization program and to each other. Results are based on 5,000 simulations from the government perspective. The four vertical dotted lines represent 0.25, 0.5, 0.75 and 1 times gross domestic product (GDP) per capita in 2024. DALY: disability-adjusted life-year, M: million, SuspVacc: Suspending vaccination.

### 2.4 Sensitivity analysis

#### 2.4.1 One-way probabilistic threshold analysis

The probabilistic threshold analysis (Figure 2 and 3) shows the threshold prices at which particular strategies become preferred based on cost-effectiveness over a range of WTP values for a DALY. For instance, from a Malawian government perspective, Figure 2A indicated that RV3-BB became the optimal strategy at WTP values (i.e., x-axis) above $125 per DALY averted \lpar &sim\semicolon \; 0.25-time GDP per capita\rpar when priced at $1 per dose or less (i.e., y-axis). This threshold price increased up to $1.6 per dose as WTP rose to $1000 per DALY averted. When the price of RV3-BB exceeds this range ($1&ndash\semicolon \; $1.6 per dose), Rotavac emerges as the optimal strategy. At low WTP values ($0&ndash\semicolon \; $60 per DALY averted), suspending vaccination remained cost-effective regardless of the RV3-BB price, consistent with the base-case findings. The duration-of-protection analysis (Figure 2B) showed that as protective durability increased, RV3-BB became cost-effective when the average duration of protection exceeded 38 weeks; below this period, the 3-dose Rotavac schedule was the optimal strategy at values above $125 per DALY averted. The switch-cost threshold analysis (Figure 3C) illustrated that the results were relatively insensitive to the one-off fixed implementation costs. The Rotarix 3-dose strategy was extendedly dominated and therefore was not shown in this analysis. It is worth noting that decision uncertainty was greatest around the lines where the preferred strategy changed.

**Figure 2:**
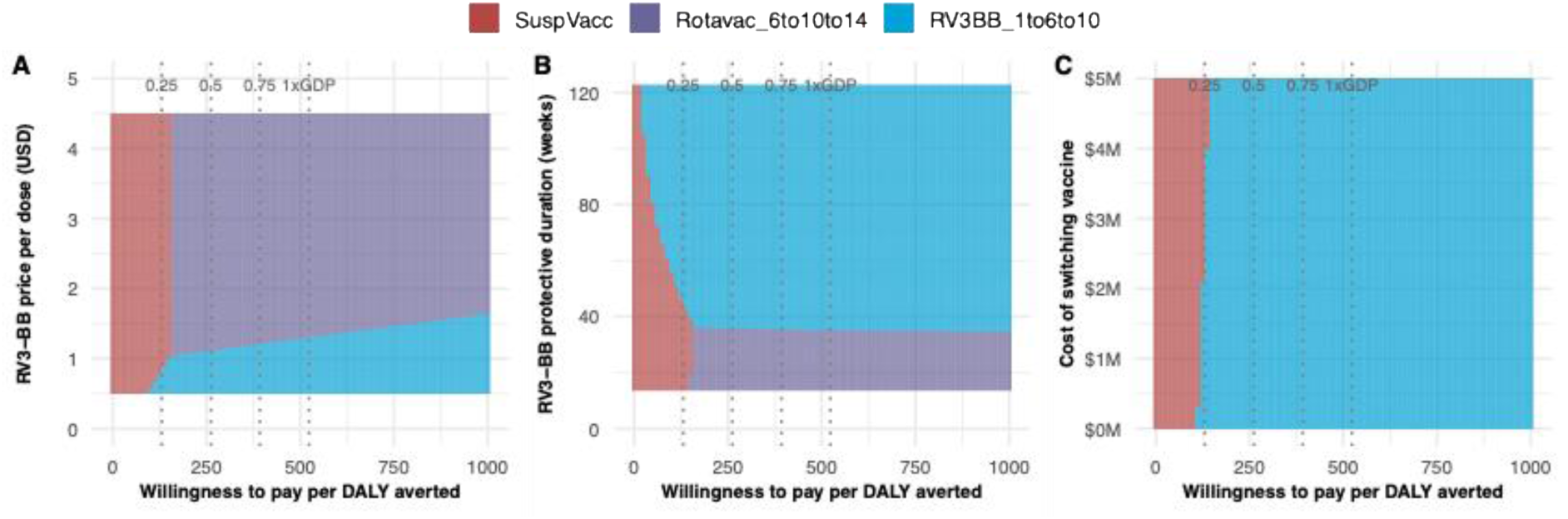
**One-way probabilistic threshold analysis in Malawi**. Four strategies were compared with the current national immunization program (Rotarix 2-dose) and to each other. Three parameters were evaluated independently: RV3-BB price (A), RV3-BB duration-of-protection (B) and vaccine program switch-cost (C). Results are based on 5,000 simulations from the government perspective. The 4 dotted lines represent 0.25, 0.5, 0.75 and 1 times gross domestic product (GDP) per capita in 2024 in Malawi. M: million, SuspVacc: Suspending vaccination, DALY: disability-adjusted life-year.

**Figure 3:**
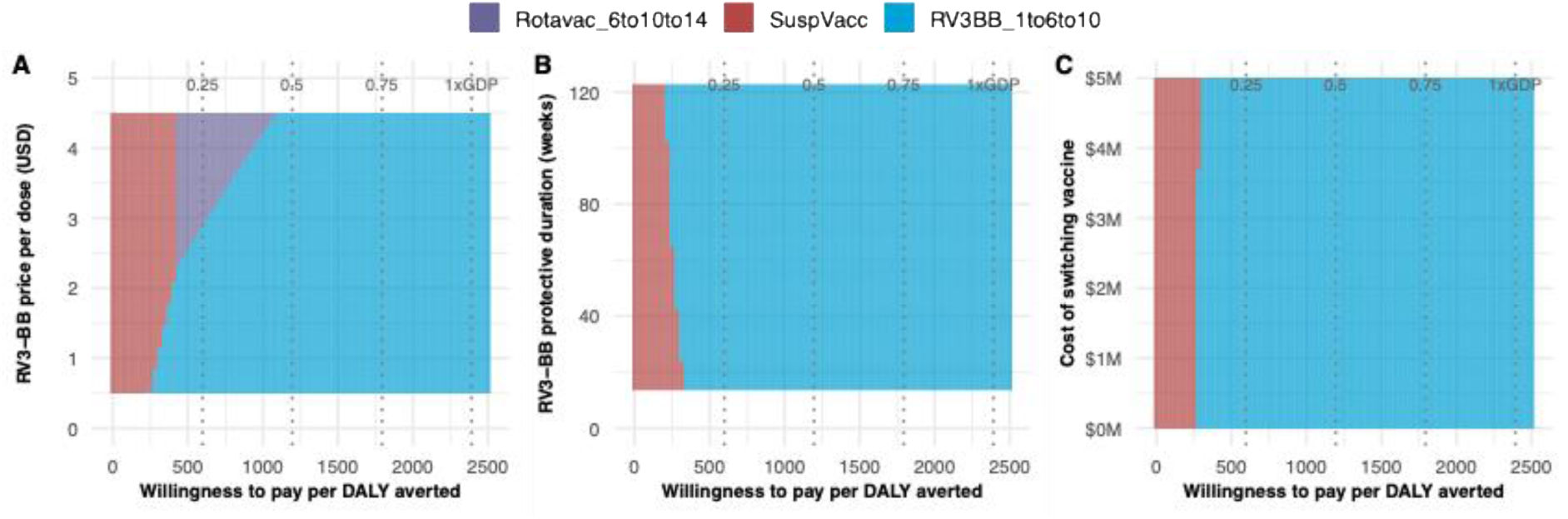
**One-way probabilistic threshold analysis in Ghana**. Two strategies were compared with the current national immunization program (Rotavac 3 dose strategy) and to each other. Three parameters were evaluated independently: RV3-BB price (A), RV3-BB duration-of-protection (B) and vaccine program switch-cost (C). Results are based on 5,000 simulations from the government perspective. The 4 dotted lines represent 0.25, 0.5, 0.75 and 1 times gross domestic product (GDP) per capita in 2024 Ghana. M: million, SuspVacc: Suspending vaccination, DALY: disability-adjusted life-year.

From the Ghanaian government perspective, the price threshold analysis (Figure 3A) indicated that RV3-BB was the optimal strategy at higher WTP values (above $1\comma \; 100 per DALY averted\comma \; less than half of the 2024 GDP per capita\rpar regardless of the vaccine price and duration of protection of RV3-BB and program switch-cost. At WTP values between $400 and $1\comma \; 100 per DALY averted\comma \; and RV3-BB priced between $2.3 and $4.5 per dose\comma \; the Rotavac strategy was optimal. At low WTP values \lpar $0–$250 per DALY averted\rpar \comma \; suspending vaccination remained optimal across all RV3-BB prices \lpar $0.5-4.5). The duration-of-protection analysis found that as protective durability increased, RV3-BB became optimal at lower WTP values, with overall limited impact (Figure 3B). Similar to Malawi, the vaccine program switch-cost also showed little impact on the results (Figure 3C).

The one-way threshold analyses from the societal perspective are shown in Figures S.6 (Malawi) and S.7 (Ghana). Findings were consistent with those from the government perspective, with RV3-BB and Rotavac becoming the preferred strategies at lower WTP values as the indirect costs are captured.

#### 2.4.2 Two-way probabilistic threshold analysis

Building on the one-way price threshold analysis, we further explored uncertainty in RV3-BB vaccine price and duration of protection using two-way threshold analyses under five WTP values from the government perspective.

In Malawi, at a theoretical WTP value of $0 per DALY averted \lpar i.e. assuming health improvements have no value to policy makers or society\rpar \comma \; RV3-BB was preferred when priced at $0.50 per dose or less, with a mean duration of protection of at least 120 weeks (Figure 4, first plot); at higher RV3-BB prices and shorter durations of protection, suspending vaccination was the optimal choice (red area). At WTP values of $250 per DALY averted and above\comma \; the optimal strategy was either Rotavac \lpar purple area\rpar or RV3-BB \lpar blue area\rpar . When RV3-BB provided more than 35 weeks of protection\comma \; it was preferred over Rotavac at a higher price per dose as values increased. The Rotarix 2- dose \lpar current NIP\rpar and 3-dose schedules were extendedly dominated due to their higher price per dose \lpar $1.79) and therefore did not appear in these plots.

**Figure 4:**
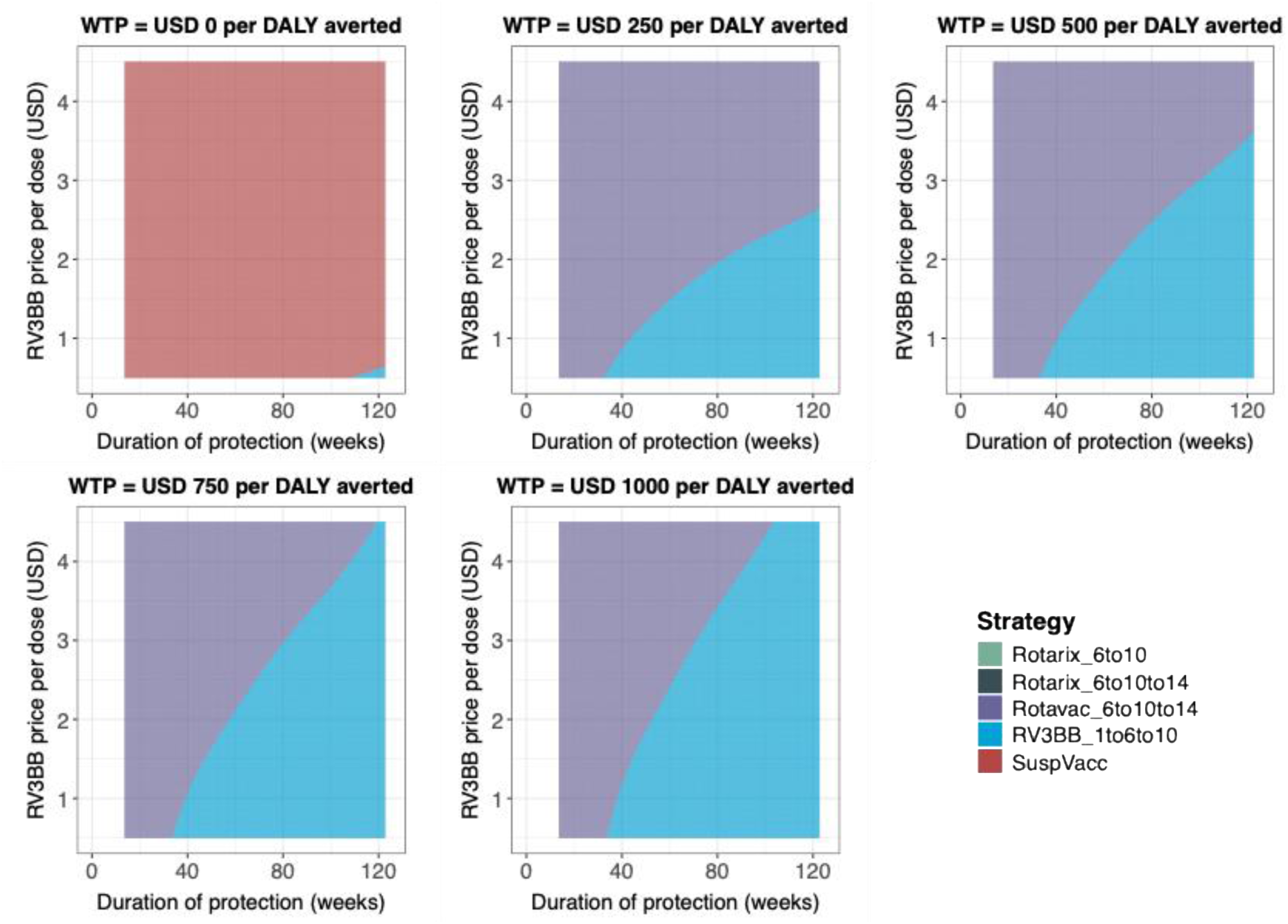
**Two-way price and duration-of-protection probabilistic threshold analysis in Malawi from the government perspective**. Four strategies were compared with the current national immunization program (Rotarix 2-dose strategy) and to each other under five willingness-to-pay values from $0 to $1000 per DALY averted. Y-axis presented price and x-axis presented duration of protection in weeks. Results are based on 5,000 simulations. SuspVacc: Suspending vaccination, DALY: disability-adjusted life-year.

In Ghana (Figure 5), at a theoretical WTP value of $0 per DALY averted\comma \; suspending vaccination remained the optimal strategy. At WTP values exceeding $625 per DALY averted, RV3-BB was preferred over Rotavac at higher price per dose alongside the increasing WTP values (increasing blue area).

**Figure 5:**
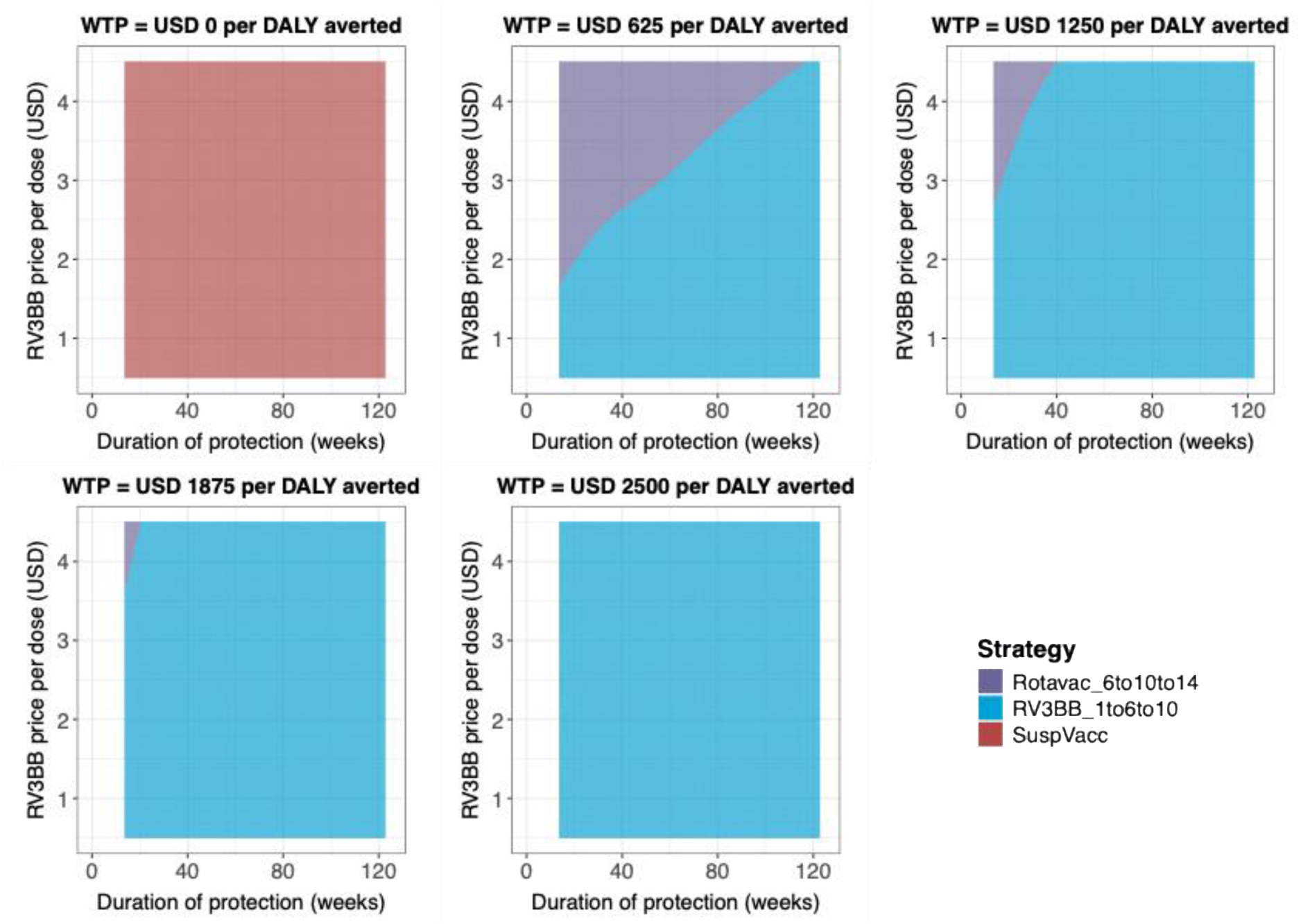
**Two-way price and duration-of-protection probabilistic threshold analysis in Ghana from the government perspective**. Two strategies were compared with the current national immunization program (Rotavac 3-dose strategy) and to each other under five willingness-to-pay values from $0 to $1000 per DALY averted. Y-axis presented price and x-axis presented duration of protection in weeks. Results are based on 5,000 simulations. SuspVacc: Suspending vaccination, DALY: disability-adjusted life-year.

The two-way threshold analyses from the societal perspective are shown in Figures S.8 (Malawi) and S.9 (Ghana). Compared with the findings from the government perspective, the color pattern generally shifted towards the lower right in both countries (i.e. RV3-BB becomes preferred at higher prices and lower durations of protection). This shift was clearly observed for Malawi at a WTP value of $0\comma \; indicating RV3-BB had a higher chance of being preferred if the average duration of protection is greater than 80 weeks and at low price per dose \lpar i.e. $0.5-1.2).

Additional two-way threshold analyses were conducted using higher ($1.15\rpar and lower \lpar $0.6) fixed prices of Rotavac in both countries from both government and societal perspectives (Figure S.10-17). Overall, the results showed that small variations in the price of Rotavac had limited impact on the preferred strategy. An exception was observed at high WTP value (around $1\comma \; 000 per DALY averted\rpar in Malawi\comma \; where the 3-dose Rotarix strategy could become preferred when RV3-BB was priced above $2 per dose and had a duration of protection of 40–80 weeks (Figure S. 10-11).It is worth noting that uncertainty increased beyond a WTP of $1,000 per DALY averted, consistent with the rising EVPI trend shown in Figure 2.

As Malawi is in the Gavi initial self-financing phase and pays a copayment of $0.20 per dose for Gavi-supported vaccines ^25^\comma \; a Malawi-specific scenario was conducted assuming both Rotarix and Rotavac prices of $0.2 per dose from the government perspective. In contrast to the base-case (Figure 4), adding a third dose of Rotarix became the optimal strategy over Rotavac (Figure S.18) across all WTP values.

#### 2.4.3 Expected Value of Partial Perfect Information (EVPPI)

The uncertainty around the delivery cost was identified as the most influential driver of decision uncertainty in both countries (Figure S.20), hence additional threshold analyses were conducted (Figures S.21 and S.22). The parameters of price and duration of protection of RV3-BB and the program switch cost were excluded from this analysis, as they had been investigated in the one- and two-way threshold analyses. In Malawi, uncertainties around hospitalization, outpatient, and non–MA episode costs ranked among the most influential parameters, whereas in Ghana, CFR-related parameters were ranked higher than cost parameters. These differences likely reflect the greater number of competing vaccination strategies in Malawi, and these strategies all offer high levels of protection against severe disease.

## 3 Discussion

Rotavirus vaccines show reduced effectiveness in LMICs, highlighting the need to explore alternative vaccination strategies to enhance vaccine performance. We therefore aimed to identify the conditions under which switching to a neonatal vaccine would be the optimal strategy in LMICs, using Malawi and Ghana as illustrative examples. We conducted a full incremental cost-effectiveness analysis of rotavirus vaccination from both government and societal perspectives, comparing all relevant strategies, including the introduction of the neonatal RV3-BB vaccine and alternative infant dosing schedules. Overall, our findings indicate that that an early birth dose of RV3-BB, administered at 1 week of life, would save lives and generate substantial public health benefits compared to the current NIP in both countries. Results indicate that either Rotavac or RV3-BB could be the optimal strategy depending on RV3-BB pricing and durability of protection at a WTP value greater than a quarter of GDP per capita.

Moreover, RV3-BB was increasingly likely to become the optimal option in both countries as WTP increased. Suspending vaccination led to a rapid increase in RVGE disease burden but was the preferred strategy at zero or very low WTP thresholds, because savings in health care did not exceed the vaccination costs. Between-country differences were also observed. Using identical WTP values, RV3-BB was more likely to be the optimal choice in Ghana than in Malawi, due to lower response rates for the second and third dose of Rotavac and short duration of protection, as well as fewer competing strategies in Ghana.

The one-way and two-way threshold analyses visually demonstrated how uncertainty in the price and duration of protection of RV3-BB would influence the choice of the optimal vaccination strategy in both countries. At WTP values above zero, RV3-BB showed an increasing probability of being the preferred strategy as WTP rose, while price and duration of protection of RV3-BB became less influential at higher WTP values, especially around 0.5–1 times GDP per capita. In Malawi, Rotavac had a higher chance of being the optimal strategy. RV3-BB would also need to be priced lower in Malawi than in Ghana to be preferred over Rotavac under the same WTP values.

Uncertainty around the choice of the cost-effective strategy plays a critical role in decision-making and must be carefully assessed. Our analysis estimated EVPI, representing the opportunity cost of making a wrong decision, to be large in both countries. Similar values of approximately $10 million were observed at the switch point between suspension of vaccination and RV3-BB. In Ghana, EVPI declined as WTP increased, reflecting reduced decision uncertainty at higher WTP levels. In contrast, EVPI increased at higher WTP values in Malawi, reflecting greater decision uncertainty driven by the competition with the 3-dose Rotarix strategy, which offers greater protection albeit at a higher price per dose. It would be valuable to conduct further studies to obtain more precise estimates of the duration of protection of RV3-BB, as this parameter plays a critical role in determining its cost-effectiveness at different price levels. Improved evidence on durability would reduce decision uncertainty and better inform policy decisions regarding the introduction of RV3-BB in LMIC settings.

Malawi and Ghana were selected as illustrative examples for the Gavi financing policy, with Malawi in the Gavi initiation phase and Ghana in the accelerated transition phase^25^. When the Gavi-negotiated prices of Rotarix ($1.79\rpar and Rotavac \lpar $0.7) were used in the base case, RV3-BB or Rotavac would be the optimal choice depending on price and durability of RV3-BB and WTP values. However, Malawi is in the Gavi initial self-financing phase, paying a $0.20 per-dose co-payment for Gavi-supported vaccines, with the remaining cost covered by Gavi ^25^. In the sensitivity analyses, we therefore applied only this copayment per dose for Rotarix and Rotavac. The results showed that RV3-BB or adding a third Rotarix dose was preferred over Rotavac, illustrating how optimal choices can vary under alternative perspectives. Alternatively, Ghana switched from 2-dose Rotarix to Rotavac in 2020, due to the lower price of Rotavac, making it more financially sustainable during the accelerated transition phase on its way to full self-financing by 2030. Our analysis showed that RV3-BB could offer additional health benefits and save direct medical costs.

From a program implementation viewpoint, our analysis evaluated the introduction of the new neonatal RV3-BB vaccine in both countries. Although adding a birth dose may require additional implementation efforts, these challenges are likely to be moderate given that RV3-BB is orally administered and that coverage of birth-dose vaccines such as BCG are high in both countries (99% in Malawi and 92% in Ghana ^26^). We applied a one-off implementation cost of approximately $1 million for introducing RV3-BB and for switching to Rotavac (in Malawi only). We also evaluated adding a third Rotarix dose at 14 weeks in Malawi; we assumed no additional implementation cost for this strategy because it uses the same vaccine product. Although this may underestimate the total cost of the 3-dose Rotarix strategy, it provides a conservative assumption for the

Rotavac and RV3-BB programs in the full incremental analysis. Our results showed that the 3-dose Rotarix program was not the optimal choice in most scenarios, except when a $0.20 co-payment per dose was assumed (Figure S.18-19). Overall, our findings indicate that one-off implementation costs had a limited impact on results over a 10-year time horizon. Nevertheless, such one-off fixed costs can impose substantial financial pressure and influence decision-making in the short-term, particularly in countries at the Gavi initial financing phase and given the financial constraints of Gavi 6.0 country-level vaccine budgets.

Our analysis included an option of suspending rotavirus vaccination, a strategy that is rarely evaluated in economic assessments. Previous published literature typically focused on comparisons with no vaccination or the current NIP. By modeling vaccination suspension, we demonstrated a rapid increase in RVGE disease burden and associated mortality. Unfortunately, suspension of rotavirus vaccination has occurred in Venezuela ^6^. Our results showed that while suspension might appear cost-effective in the 10-year horizon at zero WTP due to savings in vaccination costs, it was associated with substantial harm to population health. At relatively low WTP values (approximately 25% of GDP per capita, $125 per DALY averted in Malawi and $300 per DALY averted in Ghana), RV3-BB or Rotavac became the preferred strategy.

In the existing literature, three cost-effectiveness analyses were conducted in Malawi^27–29^, three in Ghana ^24,30,31^ and a three-country analysis, including Bangladesh, Malawi and Ghana ^32,33^. Earlier studies published between 2010 and 2016 primarily compared

Rotarix or RotaTeq with no vaccination to inform decisions on the introduction of rotavirus vaccines using a static model. Berry et al. (2010) concluded that the 2-dose Rotarix schedule was highly cost-effective in Malawi at a threshold of $75 per DALY averted ^28^. Using prospective data on RVGE-associated costs\comma \; Bar-Zeev et al. \lpar 2016\rpar found that the 2-dose Rotarix program was borderline cost-neutral from both the Malawian government and societal perspectives ^10^. Abbott et al. \lpar 2010\rpar found that\comma \; when Rotateq was priced at $1 per dose, the 3-dose program was borderline cost-neutral compared with no vaccination from the Ghanaian government perspective ^31^.

Nonvignon et al. (2018) concluded that the 2-dose Rotarix program was cost-effective in Ghana at a threshold of $332 per DALY averted from a societal perspective ^30^. Our analyses are not directly comparable with these earlier studies because suspension of vaccination is not equivalent to a baseline of no vaccination\comma \; as the protective effects of prior rotavirus vaccination wane over time. Nevertheless\comma \; our results consistently indicate that in both countries\comma \; RV3-BB would be the preferred strategy at low WTP values of \lpar i.e. $125 per DALY averted in Malawi). A three-country analysis published in 2018 and updated in 2020 compared Rotarix, Rotavac, and Rotasiil with no vaccination to inform decision-making following WHO prequalification of Rotavac and Rotasiil ^32,33^. The 2020 update concluded that Rotavac and Rotasiil were the cost-effective options across all three countries, with results highly dependent on vaccine price, level of protection, and delivery costs, which is similar to our findings. Owusu et al. (2023) explicitly evaluated the cost-effectiveness of switching from Rotarix to Rotavac in Ghana and concluded that both the 5-dose and 10-dose Rotavac presentations were cost-saving compared with Rotarix ^24^. This evidence informed our decision not to include the 2-dose Rotarix schedule in the Ghana analysis. Wenger et al. (2025) compared no vaccination, RV3-BB, and three Rotarix schedules (6–10 weeks, 6–10–14 weeks, and 6–10–40 weeks) based on output from a dynamic transmission model and concluded that RV3-BB was cost-effective at a WTP threshold of $46 per DALY averted\comma \; while the 6&ndash\semicolon \; 10&ndash\semicolon \; 40 Rotarix schedule was dominated by RV3-BB and the Rotarix 6&ndash\semicolon \; 10&ndash\semicolon \; 14 schedule ^29^. This study is the most comparable to our evaluation\comma \; although we found a higher threshold of $125 per DALY averted for RV3-BB to be cost-effective in Malawi. Beyond differences between no vaccination and suspension of vaccination, our analysis assumed a lower RV3-BB vaccine price ($0.70 vs. $1.32) but higher delivery cost ($2.02 vs. $0.58) per dose, resulting in a higher overall vaccination cost for RV3-BB. We also applied a lower Rotarix price ($1.79 vs. $1.94) in the base case, reflecting reduced Gavi-negotiated prices for all rotavirus vaccines from 2024 onward.

Our analysis has several additional strengths. First, we used a dynamic transmission model calibrated to epidemiological data from Malawi and Ghana to project RVGE burden from 2025 to 2034, fully accounting for transmission dynamics, direct and herd immunity effects of rotavirus vaccination, and dose- and country-specific response rates by vaccine product. Second, we conducted a full incremental analysis that compared all relevant vaccination strategies both against the country-specific baseline NIP and against each other. In line with the WHO guideline ^18^, comparisons limited to the current NIP may fail to capture important trade-offs among alternative schedules, particularly when more than two options exist. This was especially relevant in Malawi, where both Rotavac and RV3-BB were associated with cost savings and additional DALYs averted relative to the baseline NIP. Third, as no official WTP thresholds exist for Malawi and Ghana ^34^, we explored a wide range of WTP values to inform decision-making. Fourth, we conducted extensive sensitivity analyses to assess the robustness of our results, alongside value-of-information analyses (EVPI and EVPPI) to quantify decision uncertainty and the potential value of further research.

There are several limitations to this study. Direct and indirect costs were derived from published estimates. However, these values were originally collected in 2014 in Malawi ^27^ and 2015 in Ghana ^30^, and were adjusted to the 2025 price levels, using consumer price index corrections. The indirect costs used in our study appeared relatively similar to the direct costs, which are considerably lower than the estimates reported in more recent studies from other LMICs ^35,36^. This might underestimate the true RVGE societal costs. Moreover, although we used country-specific data wherever available, some model inputs relied on bridged evidence from other countries (i.e. Kenya ^37^) due to data gaps. Furthermore, our analysis did not explicitly evaluate the 3-dose Rotasiil strategy, primarily due to a lack of data on vaccine effectiveness. Ghana was among the first countries to implement Rotavac with available impact data. As of 2025, Rotasiil has been introduced in seven Gavi-supported settings, and real-world effectiveness data are expected to become available soon. If Rotasiil were assumed to provide protection comparable to Rotavac, its cost-effectiveness results would likely be similar. However, Rotasiil is currently supplied only as a 2-dose liquid presentation priced at $0.80 per dose\comma \; which is higher than the $0.70 frozen 5-dose presentation but lower than the $1.15 liquid 5-dose presentation of Rotavac ^8^. Future analyses incorporating upcoming effectiveness data are needed to fully assess Rotasiil. Following the previous point, while Gavi-eligible countries can select among rotavirus vaccine products, the actual implementation of Rotavac, Rotasiil, or upcoming RV3-BB is subjected to supply availability. Nevertheless, our extensive sensitivity analyses covered a wide range of plausible assumptions and provide useful insights to inform decision-making under varying circumstances.

In conclusion, this dual-country cost-effectiveness analysis compared all relevant rotavirus strategies with the current rotavirus NIP accounting for decision uncertainties related to the optimal vaccination strategy. Using a validated dynamic transmission model and country-specific data, our findings suggest that the neonatal RV3-BB vaccine can be cost-effective in Malawi and Ghana and therefore may provide similar benefit in other LMICs. Overall, our results show that the preferred rotavirus vaccination strategy based on cost-effectiveness in both countries depends on vaccine price, duration of protection, and delivery costs. It will be important for program managers should consider these aspects in their purchasing decisions.

## 4 Methods

### 4.1 Dynamic transmission model

We used an age-stratified RVGE transmission model previously calibrated and validated for both Malawi and Ghana using pre- and post-vaccination RVGE case data ^38–41^. The model structure is presented in Figure 6. It was able to reproduce observed changes in RVGE case patterns associated with COVID-19–related disruptions in Malawi and with the transition from Rotarix to Rotavac vaccine in Ghana ^42,43^. To capture the natural history of RVGE, the model assumes that newborn infants are initially protected by maternally derived antibodies and become susceptible only after the waning of this immunity. Multiple infections are permitted, with infection severity, infectiousness, and duration decreasing with each subsequent episode. The first and second infections can result in moderate-to-severe RVGE, whereas later infections are typically mild or asymptomatic. The effect of vaccination was modeled by assuming that a single vaccine dose provides temporary protection, while two doses provide additional protection equivalent to that conferred by one natural infection among vaccinated and immunized infants. Infants who respond to three doses were assumed to achieve full protection against moderate-to-severe RVGE, although they might still experience mild or asymptomatic infections.

**Figure 6:**
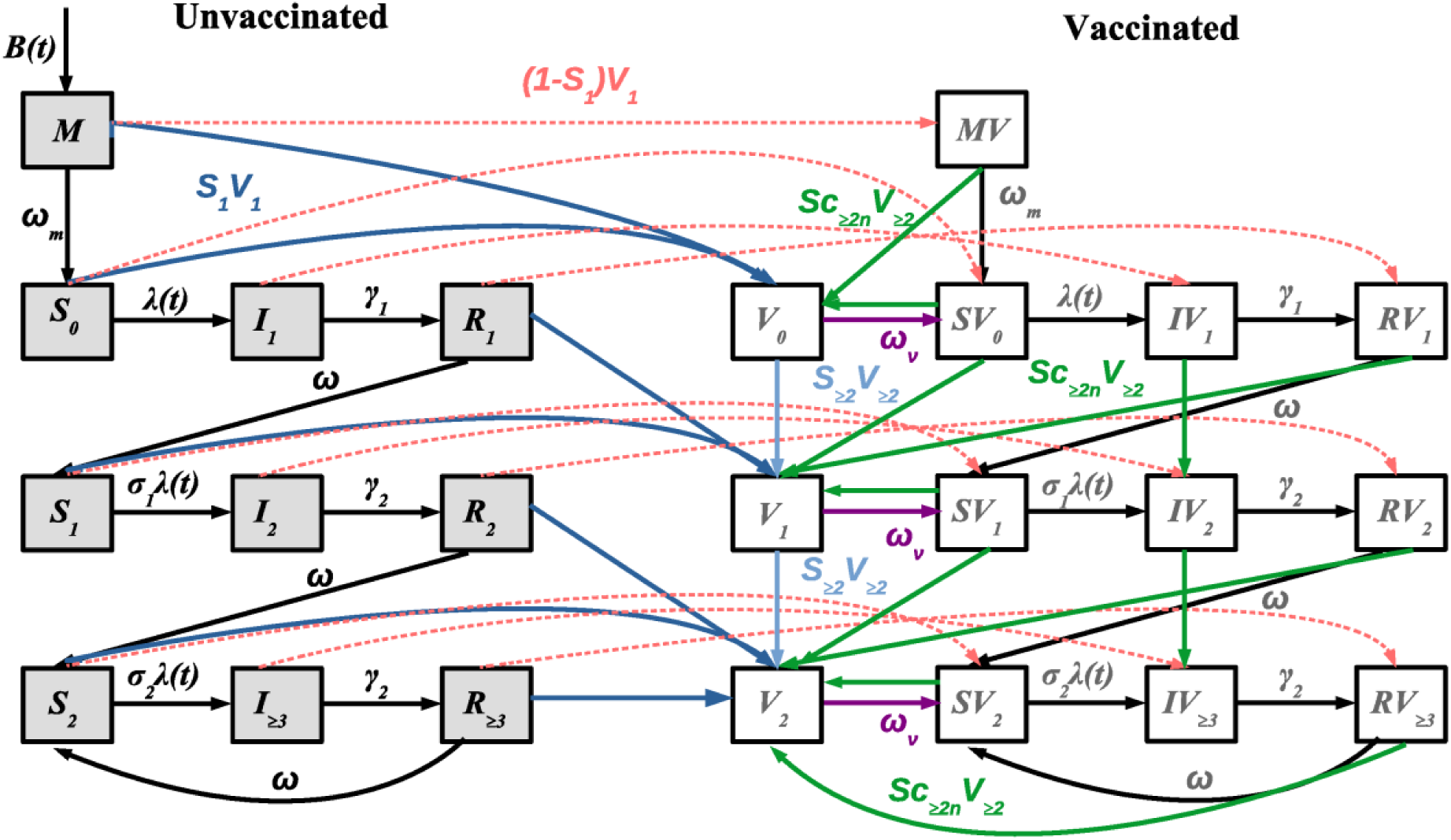
Compartmental model describing RVGE transmission. Grey and open boxes represent unvaccinated and vaccinated states, respectively, and arrows indicate the rates of movement between compartments. Dark and light blue lines denote transitions for individuals who respond to the first and subsequent vaccine doses, while red dashed lines represent transitions for individuals who do not respond to the first dose. Green lines represent the movement of individuals who fail to respond to the first dose but respond to subsequent doses, while purple lines represent the waning of vaccine-induced immunity.

As illustrated in Table 1, five vaccination strategies for Malawi and three for Ghana were modelled to estimate the number of vaccine doses, age-stratified incidence of any and moderate-to-severe RVGE over a 10-year period (2025-2034). Model outputs were stratified into 20 age groups, distinguishing annual age groups from <1 year to <5 years, 5-year age groups from 5 to 69 years, and a group aged ≥70 years. For each scenario, 5,000 samples of all model parameters except those describing vaccine effectiveness (vaccine response rates and duration of vaccine-induced immunity; see section below) were drawn from previously estimated parameters when the model was fitted to country-specific RVGE surveillance data ^39, 38 40,41^ (Tables S.1, S. 2). A detailed description of the model and parameters is provided in the Supplementary Materials.

### 4.2 Vaccine response rate and duration of protection

In both countries, we applied country-specific vaccine response rates (informed by immunogenicity data) and durations of protection. Tables S.1 and S.2 lists these model parameters for rotavirus transmission and vaccination. For Ghana, these were based on previously estimated Rotavac response rates and duration of protection ^43^. For Malawi, we used Rotarix vaccine response rates derived from a clinical trial ^44^ and the duration of protection estimated from a model fitted to RVGE surveillance data ^38^. These parameters were applied to both the 2-dose and 3-dose Rotarix schedules in Malawi. Because Rotavac is not currently used in Malawi, we doubled the variance of the Malawi-specific Rotarix duration of protection parameter for the Rotavac strategy to reflect larger uncertainty.

For RV3-BB, vaccine response rates were derived from immunogenicity estimates from a clinical trial conducted in Malawi, following the same approach used for Rotarix ^38^, and were applied to both Malawi and Ghana. Since the duration of protection for RV3-BB is unknown, we accounted for uncertainty by sampling values between the lower 95% confidence interval (CI) bound of the model-estimated duration of protection for Rotarix and the upper 95% CI bound of the duration of protection for Rotavac in both countries. Accordingly, an average duration of protection of 55.8 weeks (95% CI: 13.7–122.7 weeks) was applied in the dynamic model and examined in sensitivity analyses (see below).

### 4.3 Cost-effectiveness model

A decision tree was developed alongside the dynamic transmission model to conduct a full incremental cost-effectiveness analysis, comparing all relevant strategies. The structure of the cost-effectiveness model is in Figure 7, illustrating the care-seeking pathways for non-severe and moderate-to-severe RVGE, which were the outputs of the dynamic models. For moderate-to-severe RVGE, individuals would seek inpatient care, outpatient care, or no care, with each episode resulting in either survival or death. For non-severe RVGE, individuals may receive outpatient care or no care, and all are assumed to survive.

**Figure 7:**
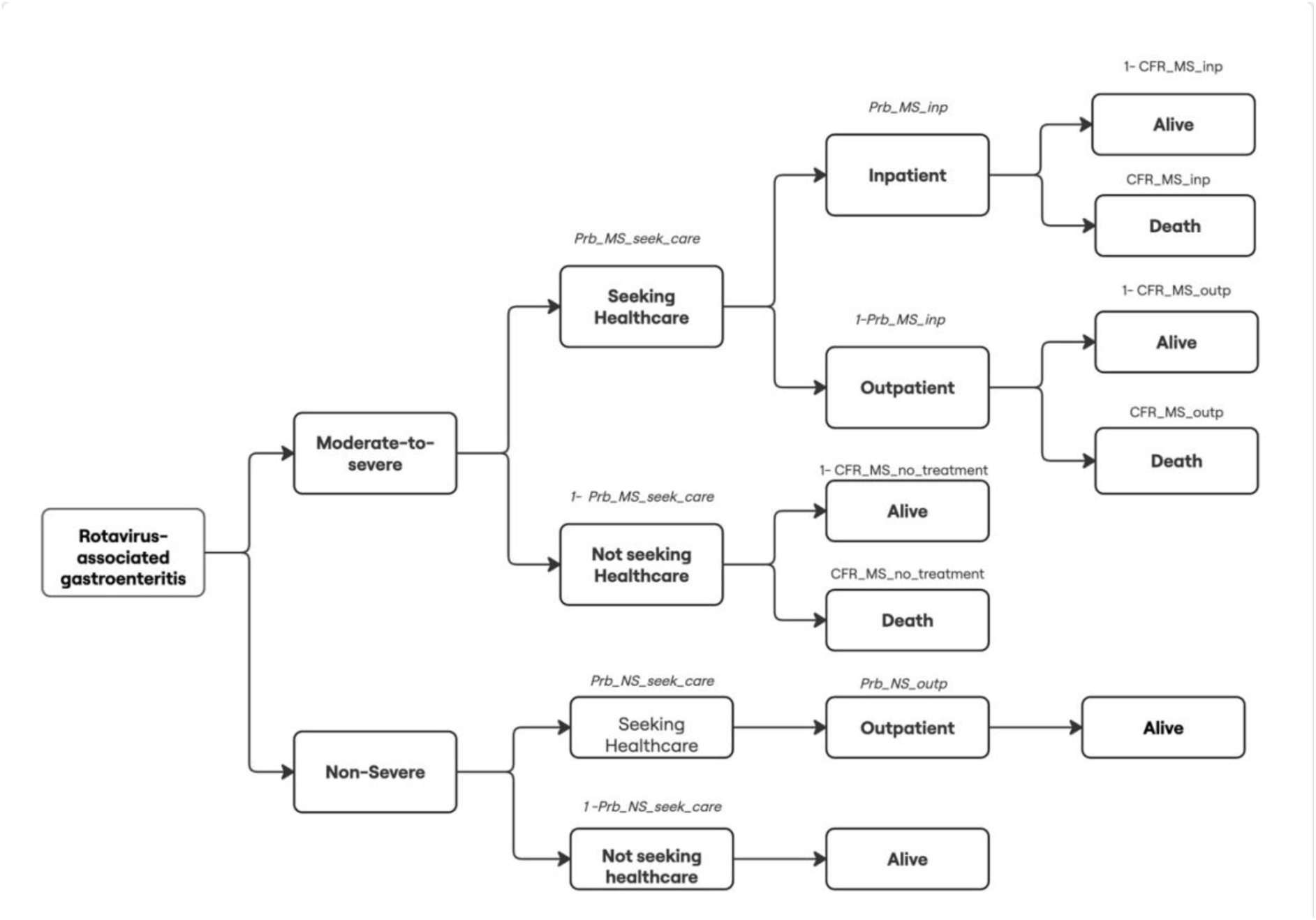
Structure of the cost-effectiveness decision-tree model. Abbreviations: Prb: probability, NS: not seeking, MS: moderate-to-severe, NS: non-severe, CFR: case-fatality ratio, inp: inpatient, outp: outpatient.

Although the dynamic transmission model included all age groups from birth to 70 years and above, the cost-effectiveness analysis focused on children under five, given the limited data on RVGE burden in older children and adults.

### 4.4 Input parameters in the cost-effectiveness model

Country-specific data were used whenever available; when unavailable, bridged data were applied. All input parameters and their associated uncertainty ranges are reported in Table 2 and Table 3.

**Table 2:** Common input parameters used in the cost-effectiveness analysis.

| Parameter<br>(notions in the model structure) | Estimate | Uncertainty Distribution | Reference/Assumption |
| --- | --- | --- | --- |
| **Health care seeking** |  |  |  |
| Probability of seeking health care for non-severe case (Prb_NS_seek_care) | 0.55 | Beta (160,133) | Non-severe diarrheal, probability of care seeking in Ghana Northern Region <sup>47</sup> |
| Probability inpatient episode for moderate-to-severe case (Prb_MS_inp) | 0.6 | Beta (1432,955) | Based on a Kenyan study <sup>37</sup> |
| Probability outpatient episode for non-severe case (Prb_NS_outpt) | 1 | Fixed | Assumption based on severity |
| **Probability of death** |  |  |  |
| CFR of inpatient episode (CFR_MS_inp) | 0.011 | Beta (7,606) | Meta-analysis of in-hospital deaths <sup>48</sup> |
| CFR of moderate-to-severe outpatient (CFR_MS_outpt) | 0.0055 | CFR_MS_inp*Unif (0,1) | Assumption: half of inpatient episode |
| CFR of moderate-to-severe no healthcare seeking (CFR_MS_no_treatment) | 0.025 | Beta (7, 259) | Meta-analysis <sup>48</sup> |
| **Indirect costs** |  |  |  |
| Indirect cost per non-healthcare seeking cases (both non-severe and moderate-to-severe cases) | 2.4 | Uniform (1.2-3.6) | Systematic review <sup>36</sup> |
| **Vaccine prices** |  |  |  |
| Rotarix price per dose | 1.79 USD | Fixed | Gavi-negotiated price <sup>8</sup> |
| Rotavac price per dose | Base case: 0.7 USD (5-dose frozen vial) | Fixed in the base case<br>Scenario analysis:<br>0.6 USD (10-dose frozen vial)<br>1.15 USD (5-dose liquid vial) | Gavi-negotiated price <sup>8</sup> |
| RV3-BB price per dose | Same as Rotavac | Fixed in the base case<br>Price threshold analysis range: \$0.5-4.5 per dose | Assumption |
| Fixed/implementation (one-off) cost of switching to a new rotavirus vaccine product/program | 1,024,365 USD | Fixed in the base case<br>Price threshold analysis range: \$0-5 million | Empirical data from Ghana <sup>24,29</sup> |
| **Vaccine wastage rate** |  |  |  |
| Single strip (i.e. Rotarix) | 0.05 | Gamma (4,80) | Vaccine wastage in Ghana, Mozambique and Pakistan <sup>52</sup> |
| 5-dose vial (i.e. Rotavac) | 0.15 | Gamma (9,60) |  |
| 10-dose vial (i.e. Rotavac) | 0.20 | Sensitivity analysis: Gamma (4,20) |  |
| **Health outcomes** |  |  |  |
| DALY weight - severe | 0.281 | Gamma (26.25, 0.11) | Global Burden of Disease study (Salomon et al 2015) <sup>54</sup> |
| DALY weight – moderate | 0.202 | Gamma (22.75 ,0.009) |  |
| DALY weight - mild | 0.061 | Gamma (17.60, 0.003) |  |
| Duration of illness to estimate DALY | 6 days | Lognormal (5.9, 6.1) | <sup>10,29</sup> |
| **Economic evaluation framework** |  |  |  |
| Discount rate | 0.03 | Fixed | WHO guideline <sup>18</sup> |

2 CFR: case fatality ratio, DALY: disability-adjusted life-year, USD: United States dollar, WHO: world health organization.

**Table 3:**
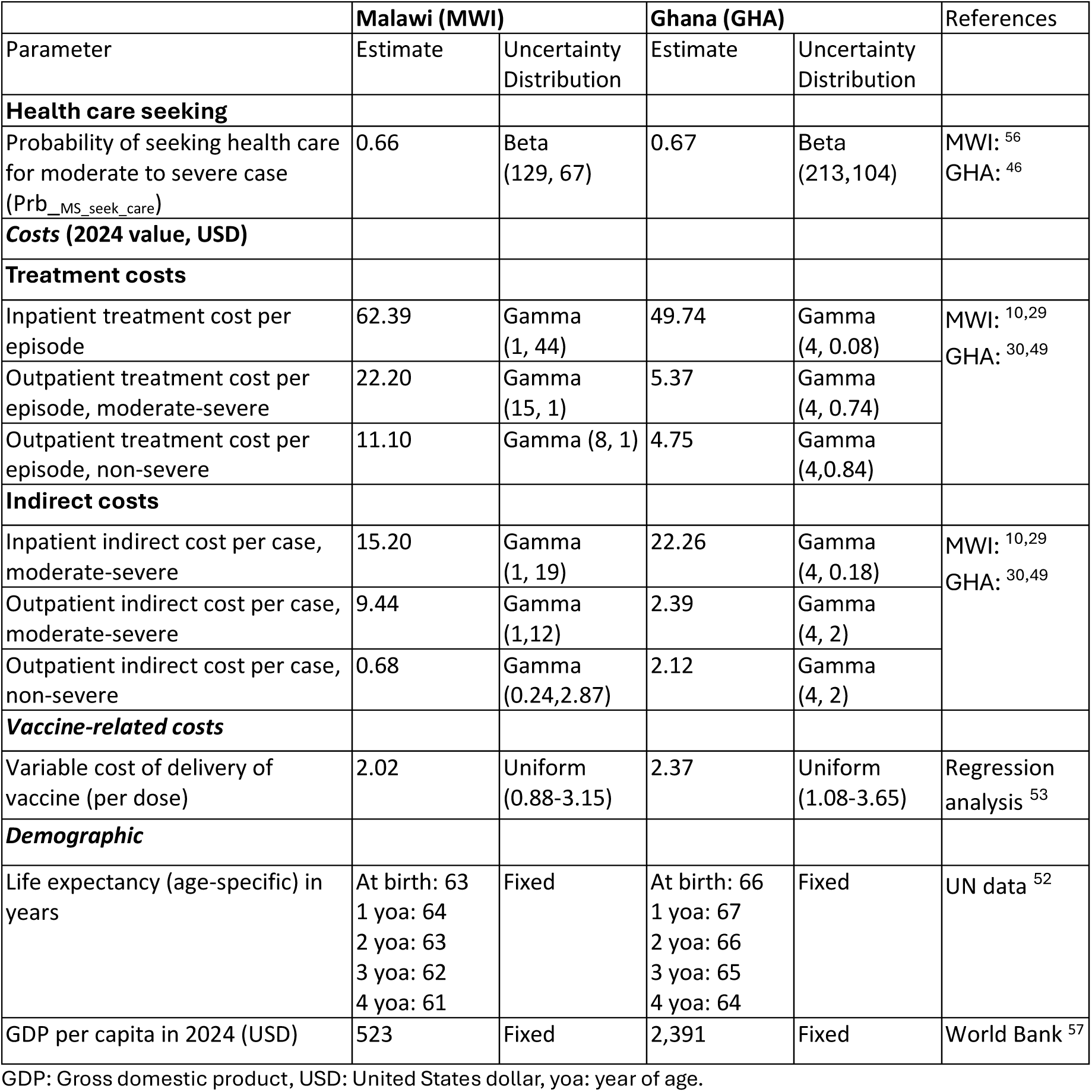
Country-specific input parameters for cost-effectiveness analysis.

#### 4.4.1 Probability of care seeking and healthcare utilization in children

Based on empirical data identified from a recent systematic review of diarrheal care-seeking ^45^ and country-specific outcomes of a regression analyses of Demographic and Health Survey data ^46^, we applied care-seeking probabilities of 67% in Ghana and 66% in Malawi for moderate-to-severe episodes, and 55% for non-severe episodes^47^ in both countries. Among moderate-to-severe cases that sought care, we assumed that 60% would receive inpatient treatment and 40% would receive outpatient care for both Malawi and Ghana, based on estimates from a Kenyan study ^37^. For non-severe cases seeking care, we assumed that all would be managed as outpatient cases. Further details are provided in Supplementary Material Section 1.2 (Tables S.3 and S.4).

#### 4.4.2 Disease-specific death

Based on a meta-analysis, we applied a case fatality ratio (CFR) of 1.1% for inpatient RVGE cases in children under 5 years of age, based on pooled hospital-based estimates for the African region ^48^. For outpatient treatment of moderate-to-severe cases, we assumed the CFR to be half that of inpatient episodes and assigned a uniform (0,1) distribution to acknowledge its large uncertainty. For moderate-to-severe cases not seeking care, a higher CFR of 2.5% was used, derived from community-based studies ^48^. For non-severe cases, we assumed 100% survival.

#### 4.4.3 Direct and indirect costs

Country-specific inpatient and outpatient direct medical costs per episode were obtained from published studies in Malawi ^10,29^ and Ghana^30,49^ from the government perspective (Table 3). From the societal perspective, household costs were also captured for both countries including direct non-medical costs and indirect costs due to productivity losses ^10,29,30^. For cases not seeking formal care (both severe and non-severe), we also included the pharmacy cost of oral rehydration solution (ORS) per episode^36^ for both countries.

All costs were converted to local currency at the year of the study, inflated to 2024 values using country-specific consumer price indices from the World Bank ^50^, and then converted back to USD using annual exchange rates ^51^.

#### 4.4.4 Vaccination related costs

The analysis applied 2025 Gavi-negotiated vaccine prices, with Rotarix priced at $1.79 per dose ^8^. Since Rotarix is supplied in 5-dose blow-fill-seal packaging\comma \; with each dose individually filled and sealed\comma \; 5&percnt\semicolon \; wastage for single-dose packaging was applied ^52^. Rotavac was available in several presentations at different prices. In our base case\comma \; a $0.70 price per dose in 5-dose frozen vials was used as base case, assuming 15% wastage due to the multiple dosage vial presentation ^52^. The 5-dose liquid presentation at a price of $1.15 and the 10-dose frozen presentation at a price of $0.6 per dose were used in sensitivity analyses ^8^. For the 10-dose vial, a higher wastage rate of 20% was assumed ^52^. Because the price of RV3-BB is unknown, we assumed it to be equivalent to the price of Rotavac in the base case ($0.70 with a 15% wastage rate). However, sensitivity analyses were conducted across a range of potential RV3-BB prices.

Country-specific costs of delivery per dose were derived from the Immunization Delivery Cost Catalogue regression analysis ^53^. As rotavirus vaccines are orally administered, we set delivery costs as the average of the lower bound of the 95% uncertainty interval (UI) and the mean estimated cost. Switching to a new vaccination schedule leads to large one-off implementation costs. Based on the cost estimated for Ghana’s transition from Rotarix to Rotavac ^24,29^, an estimated $1 million implementation cost was applied for both countries when adopting the RV3-BB strategy, or for a switch from the 2-dose Rotarix to the 3-dose Rotavac schedule in Malawi. This implementation cost was not applied when evaluating the switch from the 2-dose to the 3-dose Rotarix schedule in Malawi.

#### 4.4.5 Health outcomes

DALYs were used as the main health outcome. Years lived with disability (YLDs) were calculated by multiplying a six-day duration of illness ^10,29^ by the severity-specific disability weights for diarrheal disease from the Global Burden of Disease study ^54^. Inpatient cases were assigned the weight for severe diarrhea, outpatient and non-health seeking moderate-to-severe cases the weight for moderate diarrhea, and non-health seeking non-severe cases the weight for mild diarrhea (Table 2). Years of life lost (YLL) from RVGE premature death were calculated using country-specific life expectancy (both sexes combined) at the age of death from UN data ^52^ (Table 3).

### 4.5 Economic Analysis Framework

We conducted a full incremental cost-effectiveness analysis from both government and societal perspectives, following WHO guidelines^18^. Probabilities, costs, and health outcomes were aggregated annually by age group, and both costs and health outcomes were discounted at 3% ^18^.

#### 4.5.1 Uncertainty

To assess the impact of uncertainty in key input parameters on cost-effectiveness estimates, we calculated the EVPI and EVPPI across a range of WTP values using probabilistic sensitivity analysis ^20,21,23^. We sampled from a total of 5,000 iterations in both the transmission and the cost-effectiveness models.

The CEAC and CEAF were presented to illustrate the probability that each vaccination strategy is cost-effective across a range of WTP thresholds and to identify the preferred strategy under uncertainty.

In the absence of effectiveness data for RV3-BB in Malawi and Ghana, we hypothesized that uncertainty in its duration of protection and price would substantially influence the choice of the cost-effective vaccination strategy. To evaluate this, we applied a novel probabilistic threshold analysis, which identifies threshold values for key uncertain parameters while appropriately accounting for uncertainty in all other inputs, using a generalized additive model (GAM)-based approach ^55^.

Both one-way and two-way probabilistic threshold analyses were conducted: the one-way threshold analysis examined vaccine price, duration of protection, and switch costs independently to determine the parameter values at which the optimal strategy changes. The two-way threshold analysis can provide additional insight into how parameters jointly influence the choice of the preferred strategy.

## Declarations

### Data availability

All data generated or analyzed during this study are included in this published article. Additional data underlying the results are available from the corresponding author upon reasonable request.

### Code availability

The underlying code for this study [and training/validation datasets] is not publicly available but may be made available to qualified researchers on reasonable request to the first author (XL) and/or the senior author (VEP).

## Acknowledgments

This research was supported by a grant from National Institutes of Health/National Institute of Allergy and Infectious Diseases (R01AI112970 and R01AI137093 to VEP). The funder played no role in study design, data collection, analysis and interpretation of data, or the writing of this manuscript.

XL is also supported by a postdoctoral fellowship from the Research Foundation – Flanders (FWO) [12AF126N].

NC is a National Institute for Health and Care Research (NIHR) Senior Investigator (NIHR203756). The views expressed in this article are those of the author and not necessarily those of the NIHR, or the Department of Health and Social Care.

## Author contributions

VEP initiated the study and obtained funding. VEP and PB supervised the study. XL, EOA, PB and VEP conceptualised the study. XL analysed input data, performed a literature review and auxiliary data collection with support from CW. EOA and VEP wrote the dynamic transmission model code and calibrated the model. XL and JB wrote the cost-effectiveness model code. EOA, JK, CW, GEA, NAC, KCJ and VEP contributed to the selection of model parameters and interpretation of results for Malawi and Ghana. VEP, EOA, PB advised on intervention characteristics and scenario analyses. XL, EOA, PB and VEP wrote the initial manuscript draft. All authors critically reviewed the manuscript and provided final approval of the manuscript.

