## Supplemental file for "Cost-Effective Threshold Price for Alternative Infant and Neonatal Rotavirus Vaccines: A Dual-Country Evaluation"

### 1 Supplemental methods

#### 1.1 Dynamic transmission model

We used an age-stratified rotavirus transmission model previously calibrated and validated for both Malawi and Ghana using pre- and post-vaccination rotavirus-associated gastroenteritis (RVGE) case data [1-4] (Figure 6 in the main text).

In the absence of vaccination, rotavirus natural history was modeled as a series of transitions through maternal protection, susceptibility, infection, and recovery compartments (Figure 6). Individuals enter the model at birth into a maternally protected compartment ($M$) at a rate defined by the national birth rate ($B$). Newborns in this compartment are temporarily protected against rotavirus infection by maternally derived antibodies, which wane at a rate of $\omega_m$. Once maternal immunity has waned, individuals transition to the primary susceptible compartment ($S_0$) and can acquire a primary rotavirus infection at a rate giving by the force of infection $\lambda$. A greater proportion ($d_1$) of primary infections ($I_1$) result in moderate-to-severe RVGE, with infected individuals remaining infectious for an average duration of $1/\gamma$. After recovery, individuals move to the first recovered state ($R_1$), where they retain short-term immunity to reinfection lasting $1/\omega$ on average. Once this immunity wanes, individuals become susceptible to secondary infection ($S_1$), but at a reduced transmission rate ($\sigma_1\lambda$). Secondary infections ($I_2$) are characterized by reduced infectiousness (scaled by a factor $\rho_2$), a smaller probability ($d_2$) of developing moderate-to-severe RVGE and a shorter infectious period ($\gamma_2$) compared to primary infections. Following recovery from secondary infection, individuals enter the second recovered compartment ($R_2$) and remain temporarily immune until this protection wanes at rate ($\omega$). Following waning of this temporary acquired immunity, individuals are then transitioned into a partially immune class ($S_2$), where susceptibility to subsequent infections occurs at a further reduced rate ($\sigma_2\lambda$). Subsequent infections ($I_{\geq 3}$) are typically mild or asymptomatic ($d_3$) and have lower infectiousness (by a factor $\rho_{\geq 3}$). These individuals recover at rate ($1/\gamma_2$) and, after temporary immunity wanes ($1/\omega$), re-enter the partially immune compartment ($S_2$).

Rotavirus vaccination was modelled by introducing additional compartments that track individuals according to their vaccination and immune-response status. Transition from the unvaccinated to the vaccinated states occurred at a rate determined by country-specific first-dose vaccine coverage. Individuals who mounted an immune response to a vaccine dose (i.e. seroconverted) entered a vaccine-induced protection compartment, which conferred immunity comparable to that acquired from a single natural rotavirus infection. Those who did not respond to the first dose were assigned to a corresponding vaccinated-but-unprotected compartment. We assumed that vaccine-induced immunity wanes over time at a common rate ($\omega_v$) for all doses, after which individuals become susceptible to moderate-to-severe RVGE. Individuals who respond to subsequent doses may still experience asymptomatic or mild RVGE following the waning of vaccine-induced immunity. The model also accounted for heterogeneity in vaccine response: individuals who responded to the first dose had a higher probability of responding to subsequent doses than those who did not respond to the first dose.

*Table S. 1: Model parameters used to simulate rotavirus transmission and vaccination in Malawi and Ghana: mean and 95% uncertainty range.*

| Country-specific model parameters | Symbol | Malawi estimates | Ghana estimates |
|---|---|---|---|
| Basic reproductive number | $R_0$ | 78.8 (70.5-96.2) | 31.53 (30.36-32.78) |
| Average duration of maternal immunity | $1/\omega_m$ | 26 weeks | 1.89 (0.39-3.18) months |
| Amplitude of annual seasonality in transmission | $b_1$ | 0.174 (0.113-0.294) | 0.99 |
| Phase shift of annual seasonality | $\phi_1$ | 6.9 (4.0-11.2) weeks | 1.05 months |
| Amplitude of biannual seasonality in transmission | $b_2$ | NA | 6.63E-08 |
| Phase shift of biannual seasonality | $\phi_2$ | NA | 5.12 months |
| Reporting fraction | h | 0.017 (0.016-0.018) | 0.01 |
| Probability of responding to first vaccine dose* | $S_{C1}$ | 0.53 (0.42-0.63) | 0.811 (0.69-0.85) |
| Probability of responding to second vaccine dose (responders)* | $S_{C2}$ | 0.90 (0.72-1.0) | 0.254 (0.19-0.31) |
| Probability of responding to third vaccine dose (responders)* | $S_{C3}$ | 0.90 (0.72-1.0) | 0.154 (0.06-0.30) |
| Probability of responding to second vaccine dose (non-responders)* | $S_{C2n}$ | 0.117 (0.094-0.140) | 0.0727 (0.0243-0.1) |
| Probability of responding to third vaccine dose (non-responders)* | $S_{C3n}$ | 0.014 (0.0112-0.0168) | 0.0644 (0.0130-0.09) |
| Average duration of protection, Rotarix | $1/\omega_v$ | 45.27 (32.15-85.54) weeks | |
| Average duration of protection, Rotavac | $1/\omega_v$ | 44.92 (14.18 – 113.46) weeks | 87.58 (67.19-121.47) weeks |
| Average duration of protection, RV3-BB | $1/\omega_v$ | 55.8 (13.7-122.7) weeks | 55.8 (13.7-122.7) weeks |

*For Rotarix in Malawi, Rotavac in Ghana

*Table S. 2: Fixed model parameters used to simulate rotavirus transmission and vaccination in both countries.*

| Fixed model parameters for both countries | Symbol | Value | Source |
|---|---|---|---|
| Relative risk of second infection | $\sigma_1$ | 0.62 | [5,6] |
| Relative risk of third infection | $\sigma_2$ | 0.35 | [5,6] |
| Relative infectiousness of secondary infection | $\rho_2$ | 0.5 | [7] |
| Relative infectiousness of mild/asymptomatic infections | $\rho_{\geq 3}$ | 0.1 | [7] |
| Duration of primary infection | $1/\gamma_1$ | 1 week | [8] |
| Duration of secondary infection | $1/\gamma_2$ | 0.5 week | [9 10] |
| Duration of temporary immunity following infection | $1/\omega$ | 13 weeks | [11] |
| Proportion of primary infections that are symptomatic | $\delta_1$ | 0.41 | [5,6,12] |

| Proportion of secondary infections that are symptomatic | $\delta_2$ | 0.35 | [5,6,12] |
|---|---|---|---|
| Proportion of subsequent infections that are symptomatic | $\delta_{\geq 3}$ | 0.21 | [5,6,12] |

*For Rotarix in Malawi, Rotavac in Ghana

#### 1.2 Cost-effectiveness model

A health economic analysis plan was developed during and followed during the study; however, as it was intended to guide the analyses, it was not separately published.

##### 1.2.1 Probability of care seeking and health care utilization in children

A recent systematic review and meta-analysis by Wiens et al. (2025) [13] examined care-seeking behavior for diarrheal illness across low- and middle-income countries. Several relevant data points were identified for both Malawi and Ghana (Tables S.3 and S.4), providing empirical estimates of care utilization by severity level.

Based on these findings, we applied a care-seeking probability of 55% for non-severe RVGE in both countries based on Ghana Northen region data reported by Escribano-Ferrer et al 2016 [14]. For moderate-to-severe RVGE, we used country-specific estimates of 66% for Malawi and 67% for Ghana.

*Table S. 3: Probabilities of care seeking behavior in Malawi.*

| References | Type | Percentage (would) seek for care (numbers) |
|---|---|---|
| Lungu et al 2020 [15] | Severe diarrhea | 66% (128/194) seek care |
| Masangwi et al 2016 [16] | Diarrhea | 67% (940/1404) would seek care |
| Tebeje et al 2024 [17] | Diarrhea | Regression analysis using Demographic and Health Surveys: 66.7% |

*Table S. 4: Probabilities of care seeking behavior in Ghana.*

| References | Type | Percentage (numbers) |
|---|---|---|
| Hill et al 2003 [18] | Severe diarrhea | 61% (11/18) seek care |
| Biritwum et al 2004 [19] | Diarrhea | 22% (16/73) seek care |
| Biritwumet al 2004 [19] | Diarrhea | Tema district: 57%<br>Akwapim South district: 46% |
| | Severe diarrhea | Tema district: 60%<br>Akwapim South district: 52% |
| Krumkamp et al 2013 [20] | Acute diarrhea | 45.3% hospital attendance |
| Escribano-Ferrer et al 2016 [14] | Diarrhea | Volta Regin: 49%<br>Northern Region: 55% |
| Escribano-Ferrer et al 2017 [21] | Severe diarrhea | Volta Regin: 46%<br>Northern Region: 67% |
| Tebeje et al 2024 [17] | Diarrhea | Regression analysis: 69.2% |

### 2 Supplement Results

#### 2.1 Predicted disease and economic burden over 10 years by vaccine and schedule

The RVGE disease burden predicted by the dynamic transmission model over the 10-year period (2025–2034), stratified by vaccine product and dosing schedule, is presented in Figure S. 1.

In Malawi, compared with the current 2-dose Rotarix national immunization program (NIP), adding a third Rotarix dose at 14 weeks was associated with a 16% reduction in moderate-to-severe cases and an 8% reduction in non-severe cases. Switching to the 3-dose Rotavac schedule resulted in reductions of 14% and 7%, respectively. Implementation of the neonatal RV3-BB schedule yielded the largest impact, with reductions of 18% in moderate-to-severe cases and 11% in non-severe cases. In contrast, suspension of the rotavirus vaccination program was projected to increase moderate-to-severe and non-severe cases by 39% and 13%, respectively.

In Ghana, compared with the current NIP (3-dose Rotavac schedule), switching to RV3-BB was associated with reductions of 25% in moderate-to-severe cases and 15% in non-severe cases. In contrast, suspending the rotavirus vaccination program was projected to increase moderate-to-severe and non-severe cases by 42% and 40%, respectively.

*Figure S. 1: Projected rotavirus gastroenteritis cases in children under five from 2025–2034, by vaccination schedule and severity, in Malawi and Ghana. Results are based on 5,000 model-generated outputs from the dynamic transmission model.*

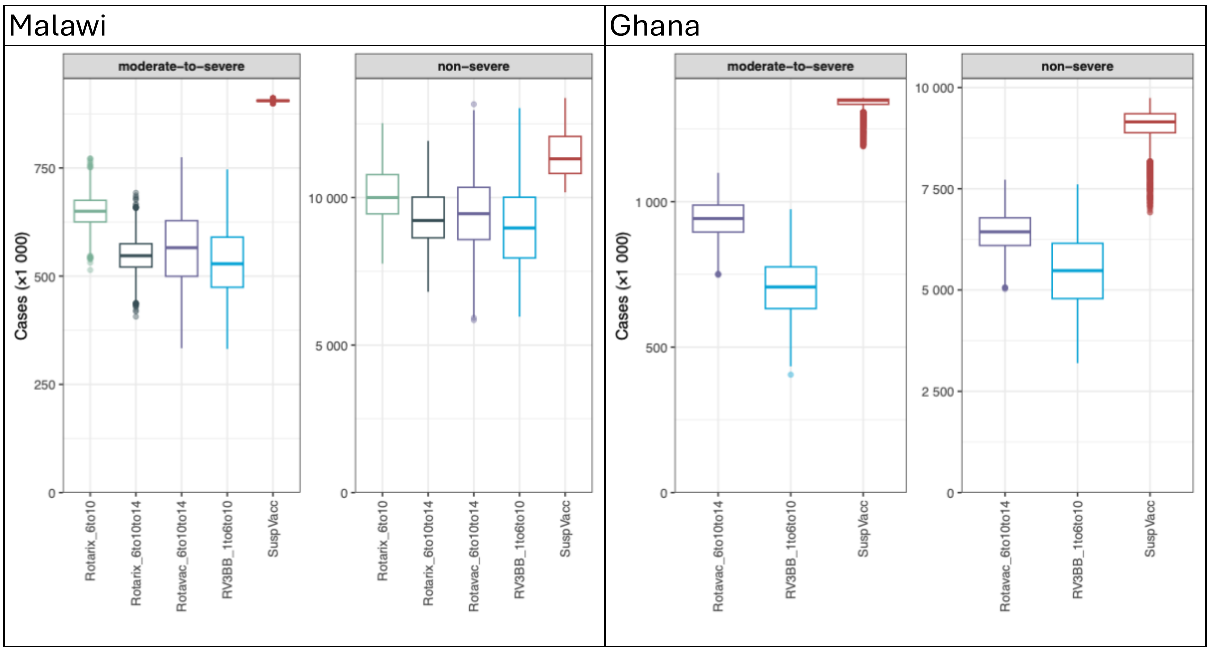

Abbreviation: SuspVacc: Suspending vaccination

The cost-effectiveness model estimated RVGE outcomes by healthcare utilization, with results reported in Table S.4.

In Malawi, versus the current NIP, introducing RV3-BB reduced RVGE-associated deaths and hospitalizations by 18%, and outpatient visits and non–medically attended (non-MA) cases by 11%. Adding a third Rotarix dose reduced deaths and hospitalizations by 16% and outpatient and non–medically attended cases by 8%, while switching to Rotavac reduced these outcomes by 14% and 7%, respectively. In contrast, suspending vaccination increased deaths and hospitalizations by 39% and outpatient and non-MA cases by 14% and 15%, respectively.

In Ghana, versus the current NIP, introducing RV3-BB reduced RVGE-associated deaths and hospitalizations by 25%, and outpatient visits and non-MA cases by 16%. In contrast, suspending vaccination increased deaths and hospitalizations by 42% and outpatient and non–MA cases by 41%.

*Table S. 5: Mean (95% credible intervals) of estimated rotavirus gastroenteritis cases in children under five, by vaccination schedule and healthcare utilization category, in Malawi and Ghana. Results are based on 5,000 simulations from the cost-effectiveness model.*

| **Malawi** | | | | | |
|---|---|---|---|---|---|
| Strategy (schedule) | Number of doses | Deaths | Hospitalizations | Outpatient | Non-MA |
| Rotarix (6,10 wk) | 10,934,673 (10,933,416 - 10,938,873) | 9,283 (4,818 - 15,320) | 257,124 (220,241 - 297,206) | 5,697,272 (4,708,832 - 6,902,622) | 4,820,264 (3,907,998 - 5,906,846) |
| Rotavac (6, 10,14 wk) | 16,400,122 (16,399,666 – 16,400,544) | 7,822 (4,044 - 12,971) | 216,684 (180,967 - 256,394) | 5,242,965 (4,241,287 - 6,462,967) | 4,429,928 (3,516,285 - 5,536,360) |
| Rotarix 3-dose (6, 10,14 wk) | 16,400,120 (16,399,658 – 16,400,543) | 8,020 (3,851 - 13,800) | 222,091 (151,232 - 286,979) | 5,313,957 (3,874,892 - 6,836,368) | 4,489,610 (3,234,758 - 5,828,697) |
| RV3-BB (1, 6, 10 wk) | 16,325,572 (16,325,427 – 16,325,714) | 7,624 (3,702 - 13,301) | 211,035 (157,067 - 273,593) | 5,078,353 (3,648,727 - 6,826,905) | 4,289,353 (3,088,405 - 5,778,025) |
| Suspending vaccination | 0 | 12,929 (6,833 - 21,048) | 358,011 (320,098 - 394,838) | 6,509,996 (5,603,569 - 7,638,224) | 5,526,779 (4,689,063 - 6,564,684) |
| **Ghana** | | | | | |
| Rotavac (6, 10,14 wk) | 24,696,379 (24,696,001 24,696,744) | 13,226 (6,784 - 21,576) | 379,734 (321,440 - 436,747) | 3,766,265 (3,128,280 - 4,419,137) | 3,230,832 (2,660,360 - 3,830,954) |
| RV3-BB (1, 6, 10 wk) | 24,723,586 (24,723,215 24,723,970) | 9,893 (4,833 - 16,923) | 283,788 (208,706 - 357,499) | 3,169,251 (2,233,418 - 4,134,853) | 2,709,270 (1,901,100 - 3,537,654) |
| Suspending vaccination | 0 | 18,797 (9,846 - 30,238) | 539,610 (487,704 - 587,082) | 22,144,018 (19,230,998 -25,105,284) | 18,421,545 (15,729,534 - 21,138,842) |

Abbreviation: Wk: weeks, MA: medically attended

The economic disease burden by vaccination schedule is shown in Figure S. 2, excluding intervention costs. In Malawi, all three alternative vaccination strategies resulted in lower discounted costs from both the government and societal perspectives, as well as fewer discounted DALYs, compared with the baseline 2-dose Rotarix schedule. Among these, RV3-BB yielded the lowest discounted costs and DALYs, although differences between the Rotavac and 3-dose Rotarix schedules were small. In contrast, suspension of vaccination resulted in substantial increases in disease burden, with total of 0.35 million discounted DALYs and approximately $96 million in direct costs and $118 million in total costs.

Similar patterns were observed in Ghana: compared with no vaccination, RV3-BB was associated with lower discounted costs and DALYs, whereas suspension of vaccination resulted in substantially higher discounted costs and DALYs.

*Figure S. 2: Discounted direct and indirect costs from the government (left panel) and societal (middle panel) perspectives, and discounted DALYs (right panel) over 2025–2034 in Malawi and Ghana. Results are based on 5,000 simulations from the cost-effectiveness model. Both costs and DALYs were discounted at 3%. Vaccination related costs were excluded.*

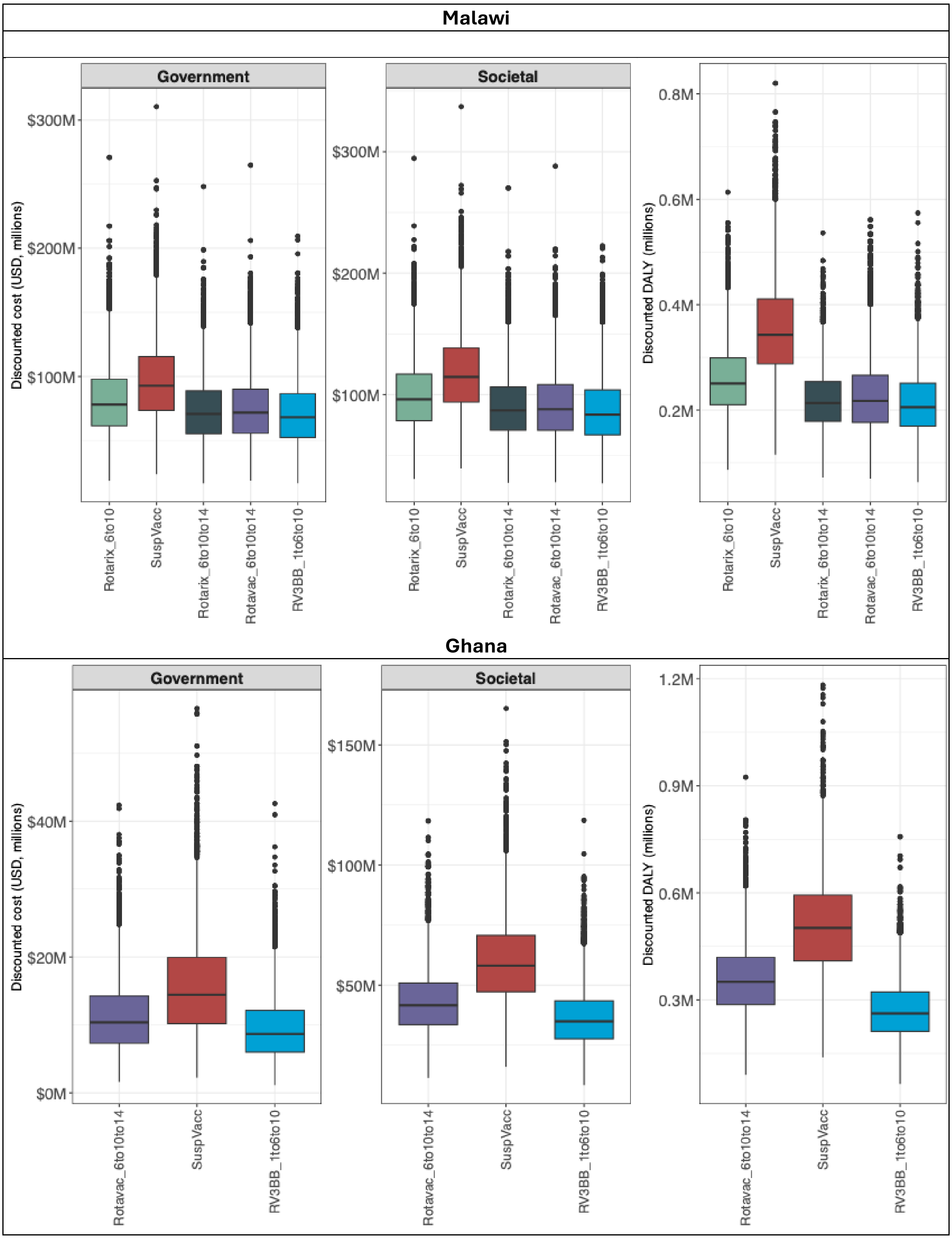

Abbreviation: DALY: disability-adjusted life-year, M: million, SuspVacc: Suspending vaccination.

Incremental results by vaccination strategy and country, including vaccination costs, are presented in Table S. 6. In Malawi, relative to the current NIP, adding a third Rotarix dose increased both direct and total RVGE-associated costs but also yielded additional DALYs averted. In contrast, switching to the 3-dose Rotavac or RV3-BB schedules resulted in cost savings and greater health gains (DALYs averted). Suspension of vaccination reduced direct and total costs compared with the current NIP but led to increased DALYs. In Ghana, switching to RV3-BB at the assumed price of $0.70 per dose (equal to Rotavac) resulted in cost savings and additional DALYs averted, whereas suspension of vaccination also reduced costs but led to increased DALYs.

*Table S. 6 Discounted incremental costs and DALYs in Malawi and Ghana. All the relevant strategies were compared with the current national immunization program and to each other. Results are based on 5,000 simulations from the government perspective. Both costs and DALYs were discounted at 3%. Intervention related costs were included.*

| Schedule | Discounted direct costs ($ million) | Discounted total costs ($ million) | Discounted DALY (thousand) | Δ costs from the government perspective ($ million) | Δ costs from a societal perspective ($ million) | Δ DALY averted (thousand) |
|---|---|---|---|---|---|---|
| Malawi | | | | | | |
| Rotarix: 6-10 (current NIP) | 119 (73-183) | 137 (88-203) | 259 (145-413) | | | |
| SuspVacc | 96 (46-168) | 118 (64-192) | 354 (198-562) | -22 (-38--4) | -19 (-35-0) | -95 (-163--47) |
| Rotarix: 6-10-14 | 130 (85-189) | 146 (98-208) | 220 (123-353) | 11 (3-19) | 9 (1-17) | 39 (21-63) |
| Rotavac: 6-10-14 | 116 (69-179) | 133 (83-200) | 225 (116-375) | -2 (-25-15) | -4 (-31-17) | 33 (-31-116) |
| RV3-BB: 1-6-10 | 113 (67-175) | 129 (80-195) | 214 (112-362) | -6 (-29-14) | -8 (-35-15) | 44 (-19-119) |
| Ghana | | | | | | |
| Rotavac: 6-10-14 (current NIP) | 80 (50-110) | 112 (75-154) | 359 (192-577) | | | |
| SuspVacc | 16 (5-33) | 60 (32-103) | 509 (276-806) | -64 (-91--37) | -51 (-82--20) | -150 (-259--72) |
| RV3-BB: 1-6-10 | 79 (50-109) | 106 (70-146) | 271 (138-455) | -1 (-6-2) | -5 (-20-5) | 87 (9-198) |

Δ: Incremental, NIP: national immunization program, DALY: disability-adjusted life-year, M: million, SuspVacc: Suspending vaccination. Red indicated negative numbers.

#### 2.2 Full incremental cost-effectiveness analysis

*Figure S. 3: Cost-effectiveness planes for Malawi (top panel) and Ghana (bottom panel) from the government perspective. The black line indicated the cost-effectiveness acceptability frontiers (CEAF). All the relevant strategies were compared with the current national immunization program and to each other. Results are based on 5,000 simulations.*

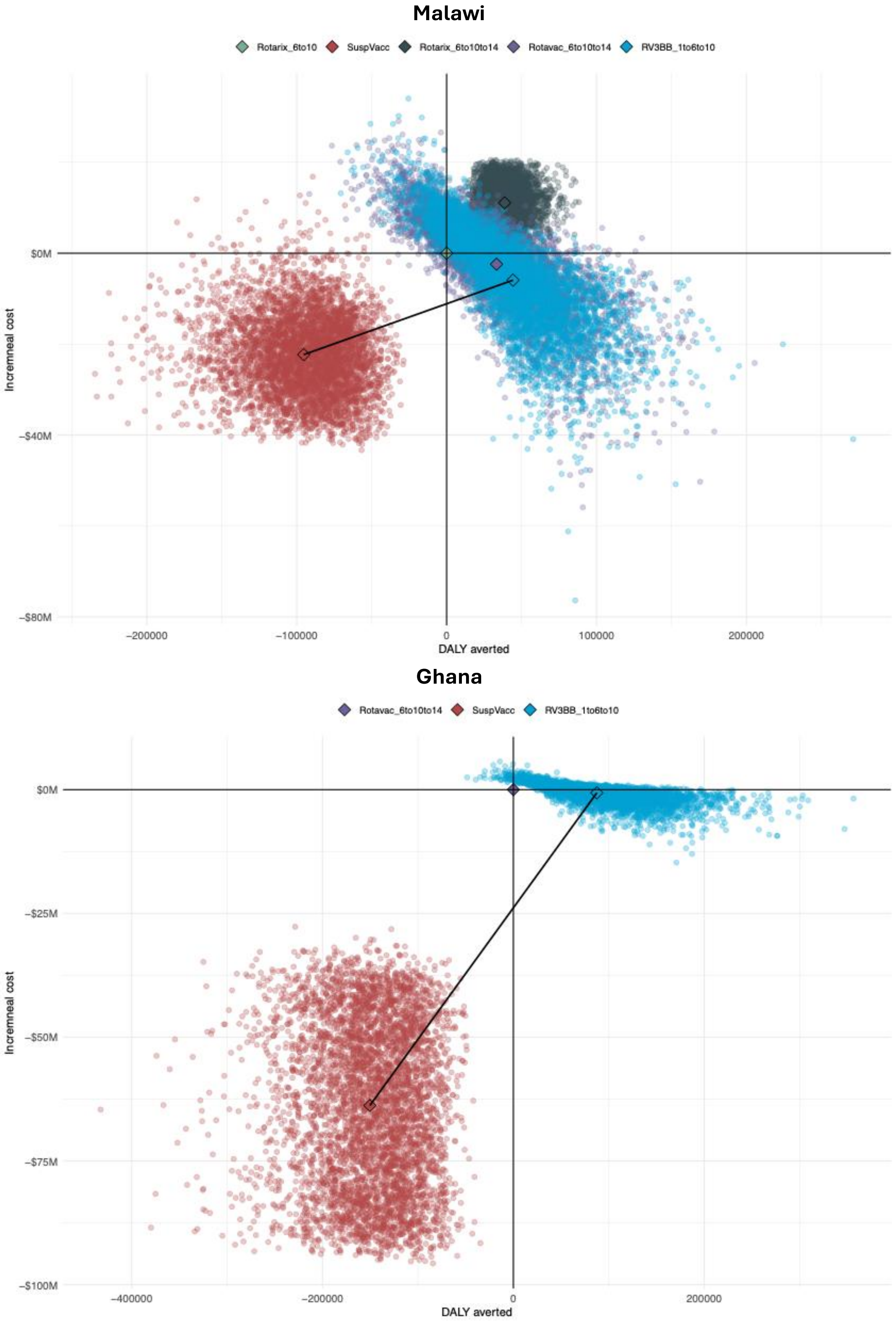

*Figure S. 4: Cost-effectiveness planes for Malawi (top panel) and Ghana (bottom panel) from a societal perspective. The black line indicated the cost-effectiveness acceptability frontiers (CEAF). All the relevant strategies were compared with the current national immunization program and to each other. Results are based on 5,000 simulations.*

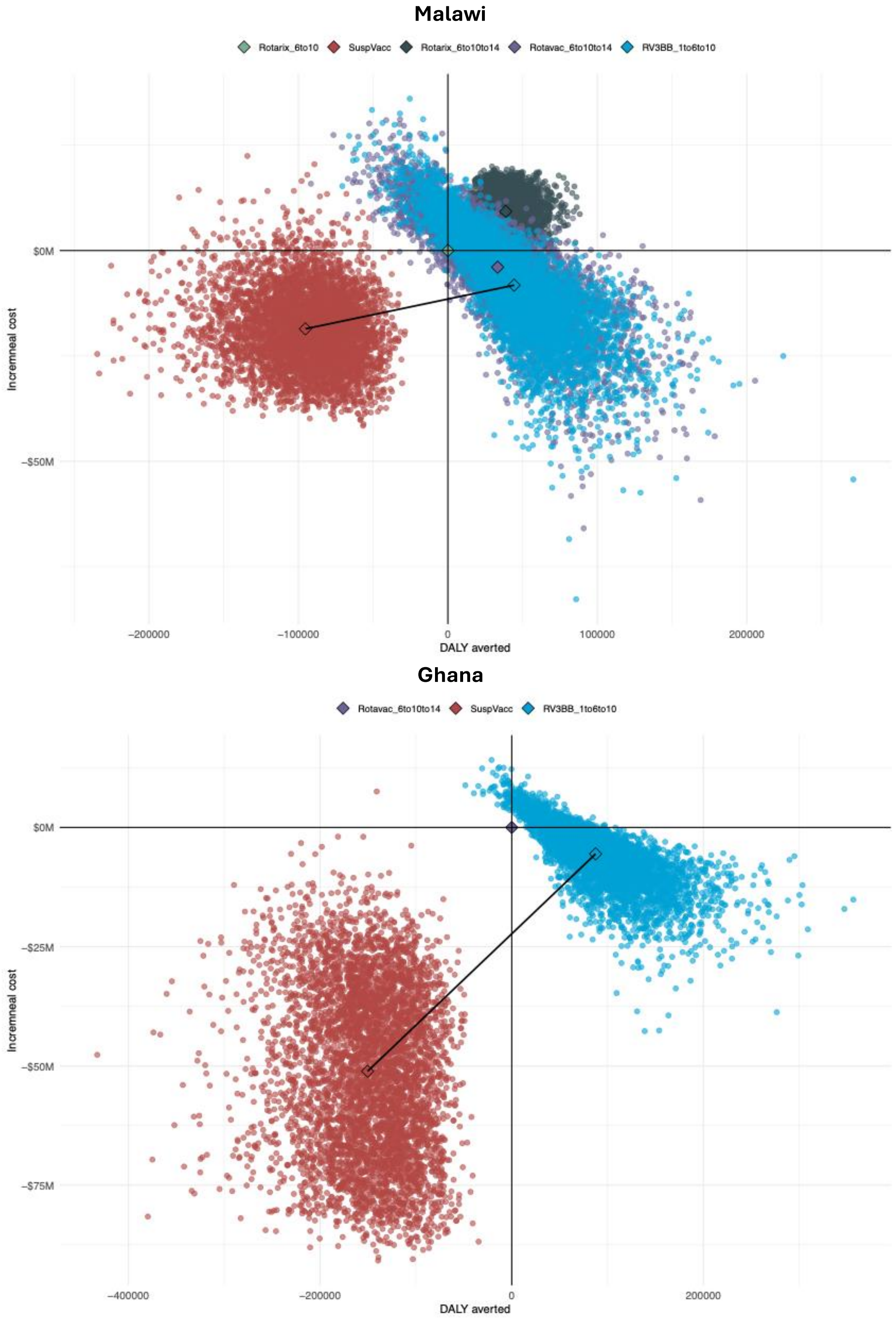

*Table S. 7: Percentage of incremental cost-effectiveness ratio (ICER) iterations by cost-effectiveness plane quadrant (N=5,000 per strategy)*

| **Strategy (schedule)** | **Malawi** | | | | **Ghana** | | | |
|---|---|---|---|---|---|---|---|---|
| | north-east | south-east | south-west | north-west | north-east | south-east | south-west | north-west |
| **Government perspective** | | | | | | | | |
| Rotavac (6, 10,14 wk) | 24.82% | 55.68% | 0.68% | 18.82% | | | | |
| Rotarix 3-dose (6, 10,14 wk) | 99.40% | 0.60% | 0% | 0% | | | | |
| RV3-BB (1, 6, 10 wk) | 21.64% | 67.72% | 0.08% | 10.56% | 42.36% | 56.36% | 0% | 1.28% |
| Suspending vaccination | 0% | 0% | 98.96% | 1.04% | 0% | 0% | 100% | 0% |
| **Societal perspective** | | | | | | | | |
| Rotavac (6, 10,14 wk) | 21.36% | 59.14% | 0.70% | 18.80% | | | | |
| Rotarix 3-dose (6, 10,14 wk) | 98.42% | 0% | 0% | 1.58% | | | | |
| RV3-BB (1, 6, 10 wk) | 18.78% | 70.58% | 0.02% | 10.62% | 19.52% | 79.20% | 0% | 1.28% |
| Suspending vaccination | 0% | 0% | 97.30% | 2.70% | 0% | 0% | 99.98% | 0.02% |

Figure S. 5 presents the cost-effectiveness acceptability curves (CEAC), cost-effectiveness acceptability frontiers (CEAF), expected net losses (ENL), and expected value of information (EVPI) for Malawi (top panel) and Ghana (bottom panel) from a societal perspective. Willingness-to-pay (WTP) thresholds ranged from $0 to $1,000 per DALY averted in Malawi and from $0 to $2,500 per DALY averted in Ghana. Compared with the findings from the governmental perspective, RV3-BB became the preferred strategy at slightly lower WTP values.

*Figure S. 5: Cost-effectiveness acceptability curve, cost-effectiveness acceptability frontier, expected net losses and expected value of information for Malawi (top panel) and Ghana (bottom panel). All the relevant strategies were compared with the current national immunization program and to each other. Results are based on 5,000 simulations from a societal perspective.*

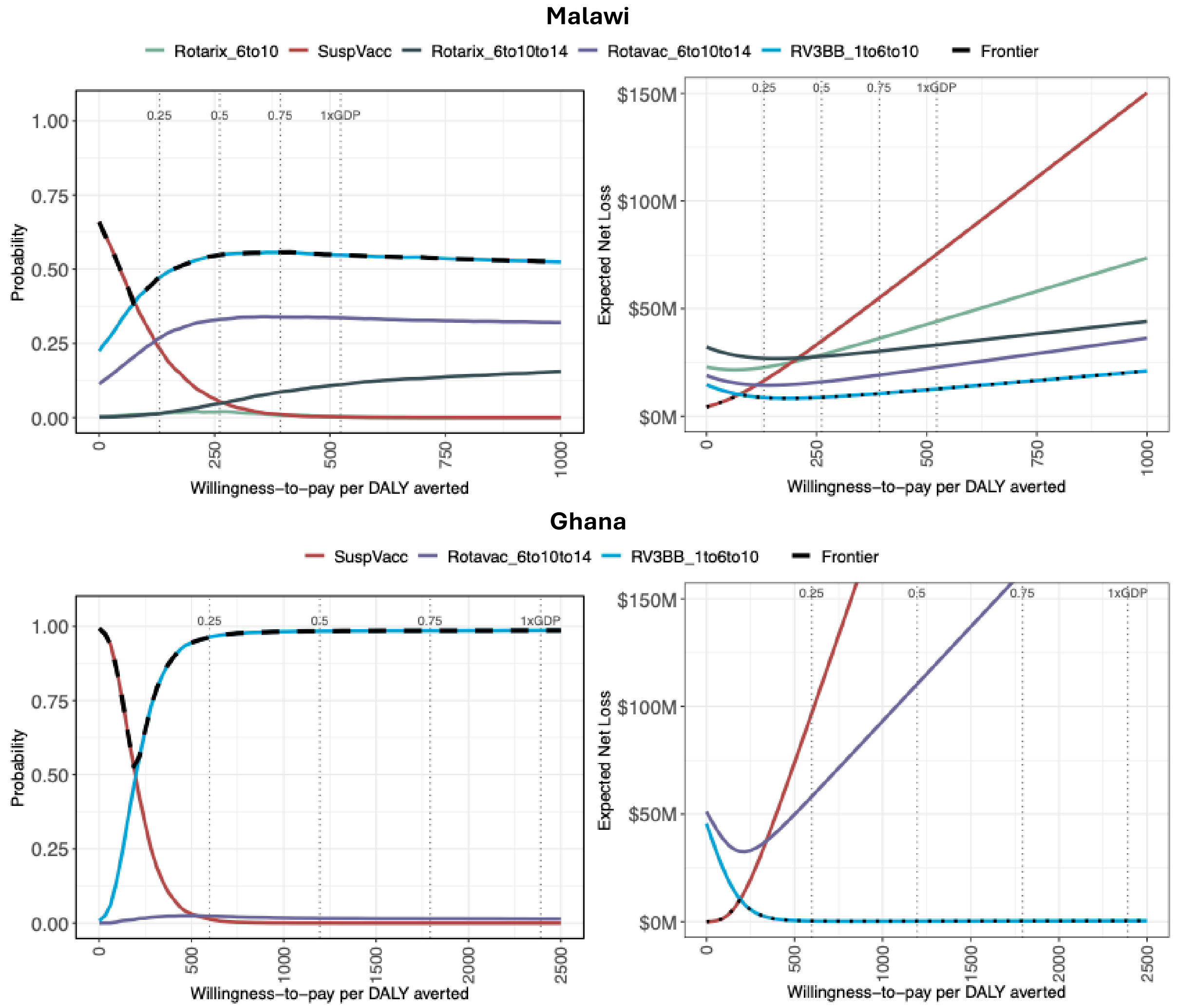

The 4 dotted lines represented: 0.25, 0.5, 0.75 and 1 times Gross Domestic Product (GDP) per capita.
Abbreviations: DALY: disability-adjusted life-year, M: million, SuspVacc: Suspending vaccination.

#### 2.3 Uncertainty and probabilistic analysis

##### 2.3.1 One-way probabilistic threshold analysis from the societal perspective

The one-way threshold analyses from the societal perspective are presented in Figure S. 6 for Malawi and Figure S. 7 for Ghana. Overall, the results were consistent with those from the government perspective (main text Figure 1 and 2), but with RV3-BB and Rotavac becoming the preferred strategies at lower WTP values, reflecting the inclusion of broader indirect costs.

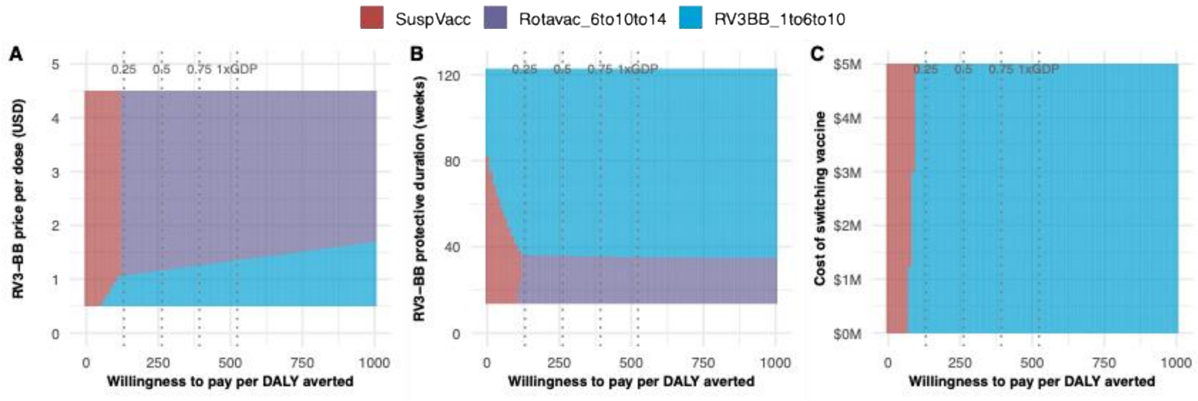

***Figure S. 6: One-way probabilistic threshold analysis in Malawi from a societal perspective**. Four strategies were compared with the current national immunization program (Rotarix 2-dose strategy) and to each other. Three parameters were evaluated independently: price (A), duration of protection (B) and switch cost (C). Results are based on 5,000 simulations.*

The 4 dotted vertical lines represented: 0.25, 0.5, 0.75 and 1 times Gross Domestic Product (GDP) per capita. Abbreviations: M: million, SuspVacc: Suspending vaccination.

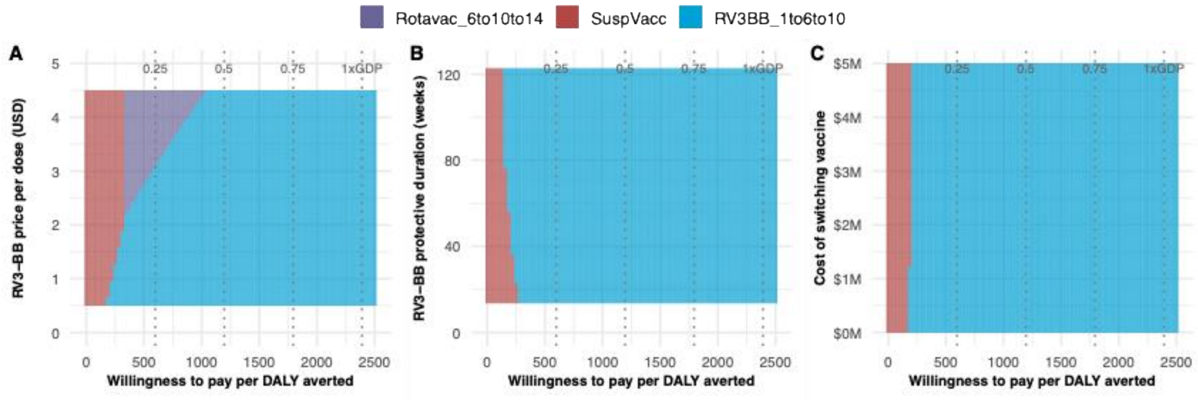

***Figure S. 7: One-way probabilistic threshold analysis in Ghana from a societal perspective**. Two strategies were compared with the current national immunization program (Rotavac 3-dose strategy) and to each other. Three parameters were evaluated independently: price (A), duration of protection (B) and switch cost (C). Results are based on 5,000 simulations.*

The 4 dotted vertical lines represented: 0.25, 0.5, 0.75 and 1 times Gross Domestic Product (GDP) per capita. Abbreviations: M: million, SuspVacc: Suspending vaccination.

##### 2.3.2 Two-way threshold analysis from societal perspective

The two-way threshold analyses examining vaccine price and duration of protection from the societal perspective are presented in Figure S. 8 for Malawi and Figure S. 9 for Ghana. Compared to the government perspective, the color pattern of the preferred strategy shifted up and to the left overall, reflecting the broader societal costs averted by vaccination. This shift was most evident in Malawi at a value of $0 per DALY averted, where RV3-BB showed a higher probability of being the preferred strategy when protection exceeded 80 weeks at lower price levels (approximately $0.50–$1.20 per dose).

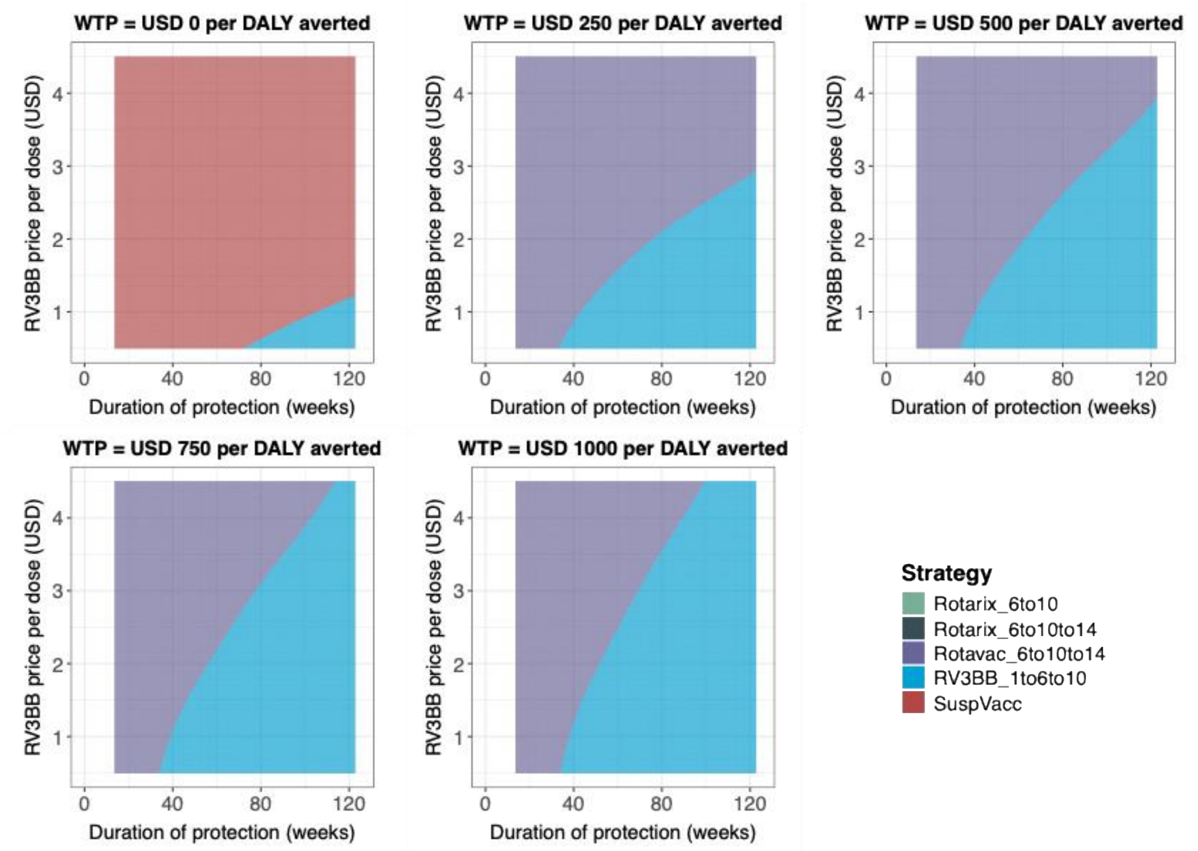

***Figure S. 8: Two-way price and duration of protection probabilistic threshold analysis in Malawi from a societal perspective****. Four strategies were compared with the current national immunization program and to each other under five willingness-to-pay values from $0 to $1000 per DALY averted. Y-axis presented price and x-axis presented duration of protection in weeks. Results are based on 5,000 simulations.*

Abbreviations: SuspVacc: Suspending vaccination, DALY: disability-adjusted life-year.

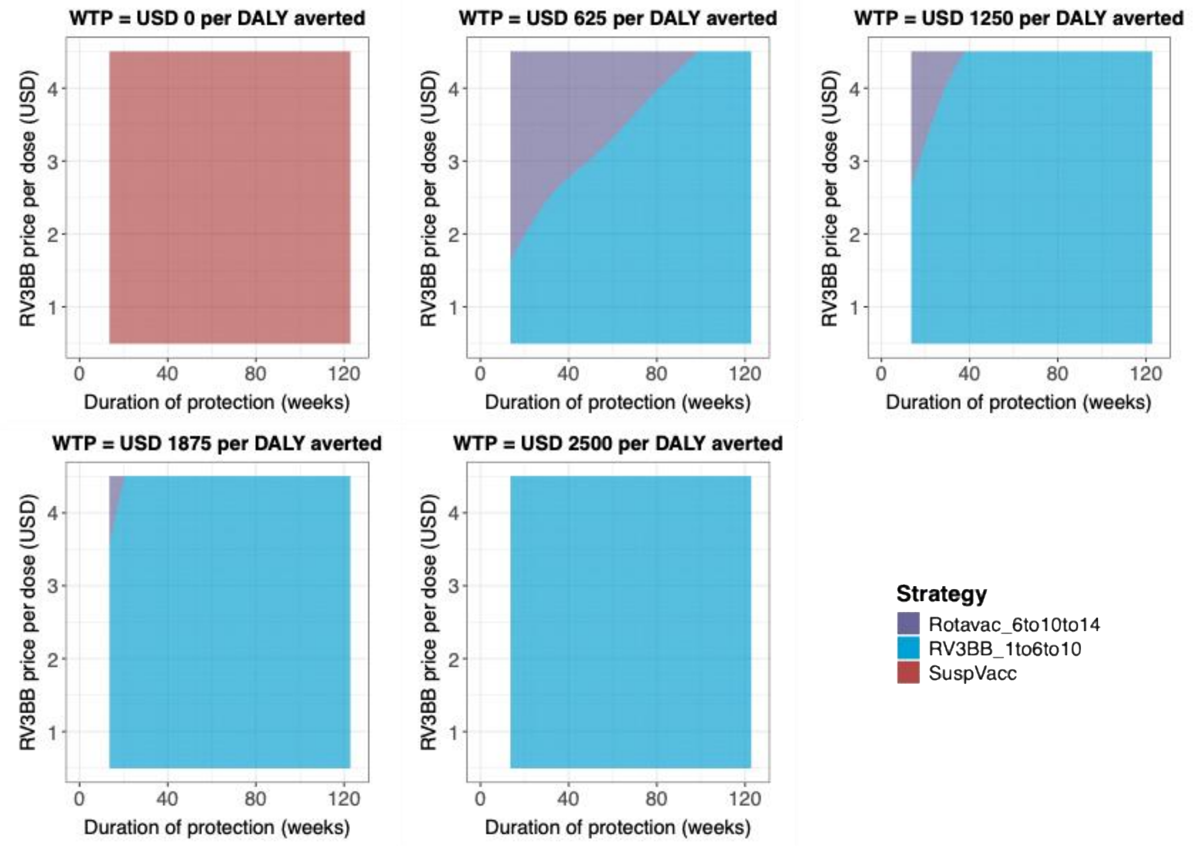

***Figure S. 9: Two-way price and duration of protection probabilistic threshold analysis in Ghana from a societal perspective****. Two strategies were compared with the current national immunization program and to each other under five willingness-to-pay values from $0 to $1000 per DALY averted. Y-axis presented price and x-axis presented duration of protection in weeks. Results are based on 5,000 simulations.*

Abbreviations: SuspVacc: Suspending vaccination, DALY: disability-adjusted life-year.

##### 2.3.3 Two-way threshold analysis when assuming Rotavac at higher Gavi-negotiated price of $1.15 per dose

Sensitivity analyses were conducted on the price of Rotavac. Assuming the 5-dose liquid vial was used in the NIP in both countries instead of the 5-dose frozen vial in the base case, a price of $1.15 per dose was applied with the same wastage rate (15%).

In Malawi, at the higher Rotavac price, results were comparable with the base case analysis (assuming $0.70 per dose for Rotavac) from both perspectives of government (Figure S. 10) and society (Figure S. 11). However, at a value of $1,000 per DALY averted, the 3-dose Rotarix schedule emerged as the preferred strategy when RV3-BB was priced above $2 per dose and its duration of protection ranged between 40 and 80 weeks (dark green area).

In Ghana, assuming a higher Rotavac price per dose, Figure S.12 and Figure S. 13 illustrate that the color pattern shifted toward the upper left at WTP values above $0 per

DALY averted compared with the base case (Figures 5 and Figure S. 9). This shift indicates that RV3-BB became the preferred strategy at a higher price when Rotavac was priced higher; however, this change had limited impact on the overall choice of the preferred strategy across all five WTP values.

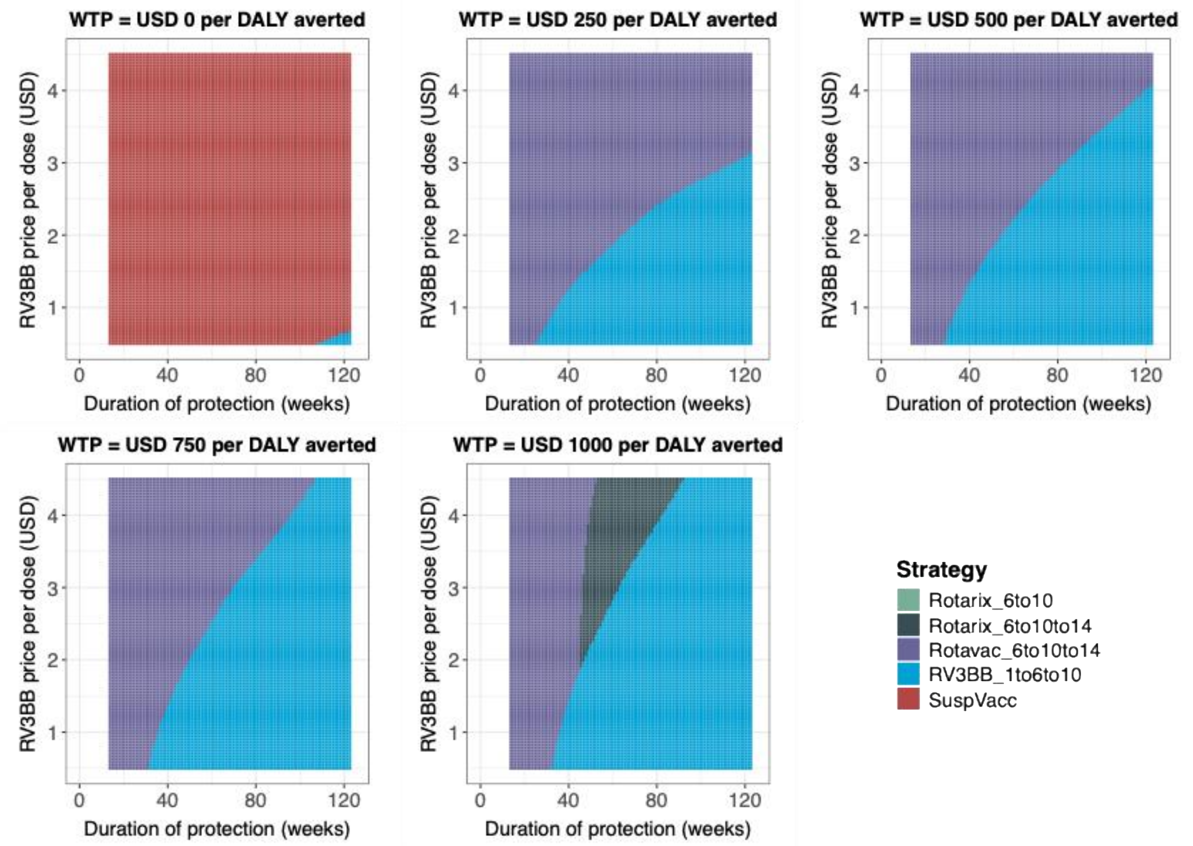

***Figure S. 10: Two-way price and duration of protection probabilistic threshold analysis in Malawi from the government perspective assuming the price of Rotavac is $1.15 per dose with 15% wastage****. Four strategies were compared with the current national immunization program and to each other under five willingness-to-pay values from $0 to $1000 per DALY averted. Y-axis presented price and x-axis presented duration of protection in weeks. Results are based on 5,000 simulations.*

Abbreviations: SuspVacc: Suspending vaccination, DALY: disability-adjusted life-year.

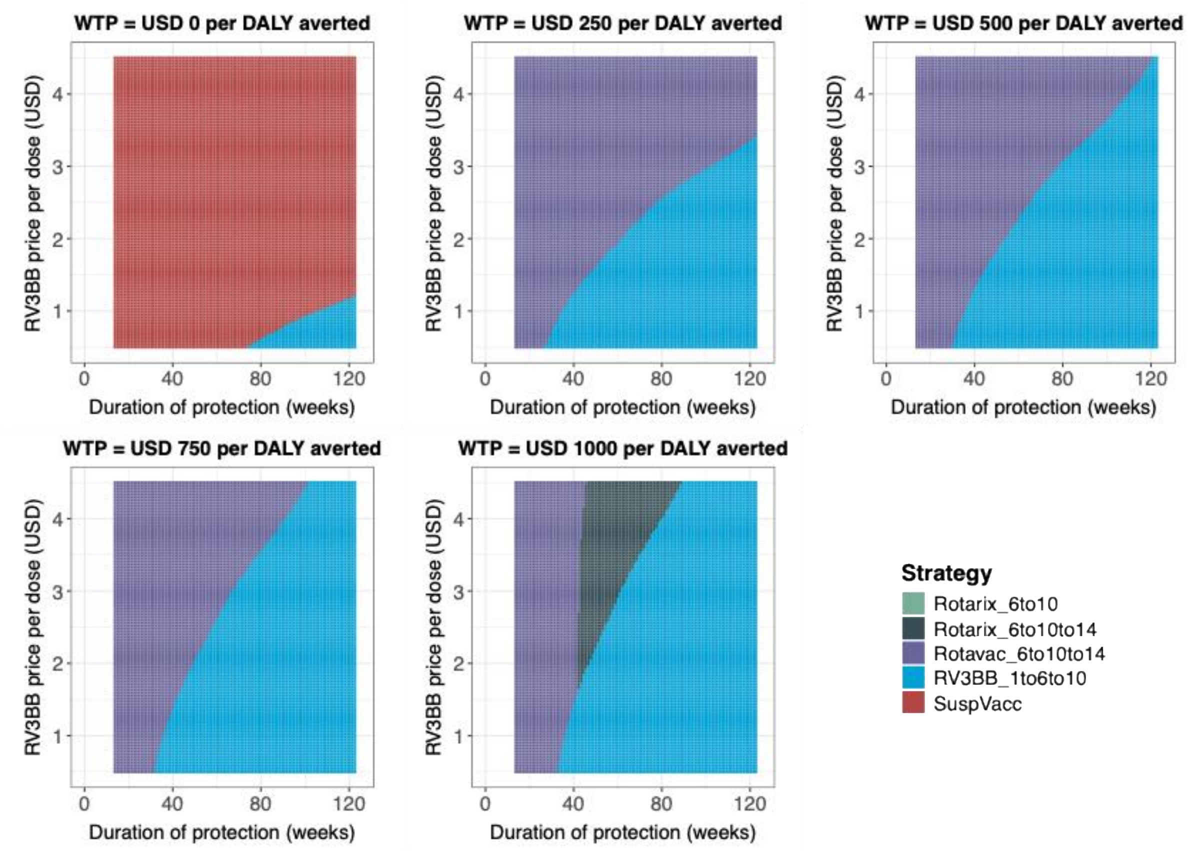

***Figure S. 11: Two-way price and duration of protection probabilistic threshold analysis in Malawi from a societal perspective assuming the price of Rotavac is $1.15 per dose with 15% wastage.*** *Four strategies were compared with the current national immunization program and to each other under five willingness-to-pay values from $0 to $1000 per DALY averted. Y-axis presented price and x-axis presented duration of protection in weeks. Results are based on 5,000 simulations.*

Abbreviations: SuspVacc: Suspending vaccination, DALY: disability-adjusted life-year.

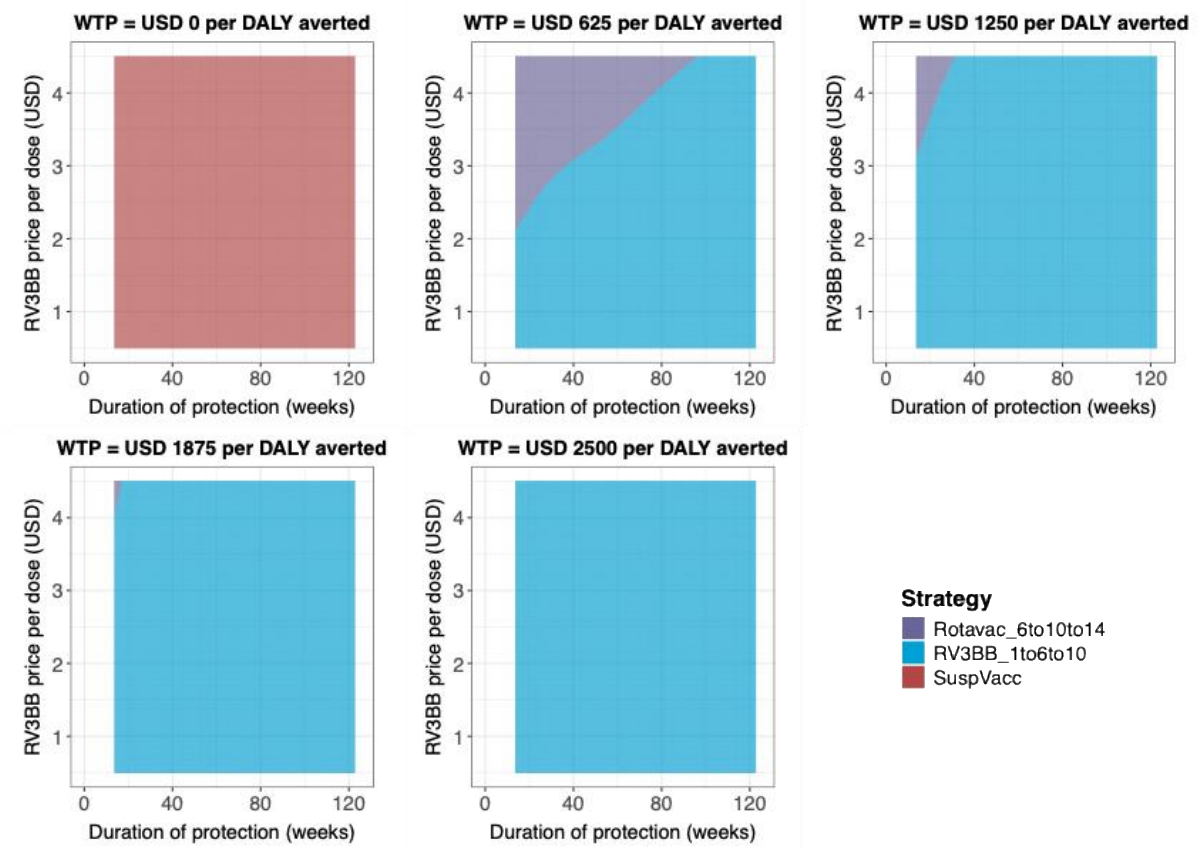

***Figure S.12: Two-way price and duration of protection probabilistic threshold analysis in Ghana from the government perspective assuming the price of Rotavac is $1.15 per dose with 15% wastage****. Two strategies were compared with the current national immunization program and to each other under five willingness-to-pay values from $0 to $1000 per DALY averted. Y-axis presented price and x-axis presented duration of protection in weeks. Results are based on 5,000 simulations.*

Abbreviations: SuspVacc: Suspending vaccination, DALY: disability-adjusted life-year.

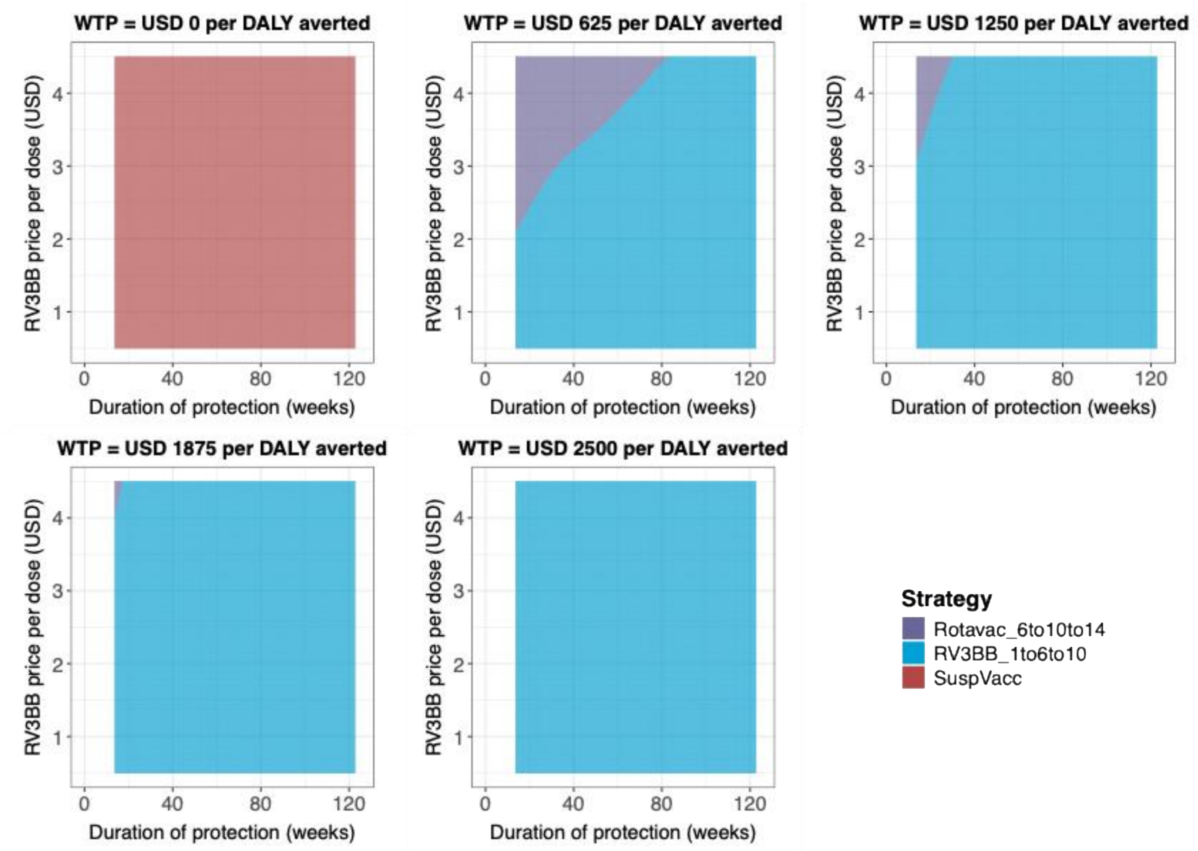

***Figure S. 13: Two-way price and duration of protection probabilistic threshold analysis in Ghana from a societal perspective assuming the price of Rotavac is $1.15 per dose with 15% wastage.** Two strategies were compared with the current national immunization program and to each other under five willingness-to-pay values from $0 to $1000 per DALY averted. Y-axis presented price and x-axis presented duration of protection in weeks. Results are based on 5,000 simulations.*

Abbreviation: SuspVacc: Suspending vaccination, DALY: disability-adjusted life-year.

##### 2.3.4 Two-way threshold analysis when assuming Rotavac at lower Gavi-negotiated price of $0.6 per dose

In this sensitivity analysis, we assumed the 10-dose frozen vial of Rotavac would be used in the NIP in both countries. The price was $0.6 per dose with a higher wastage rate of 20%.

In Malawi, at lower vaccine prices, results were comparable with the base case analysis from both perspectives (Figure S. 14 and

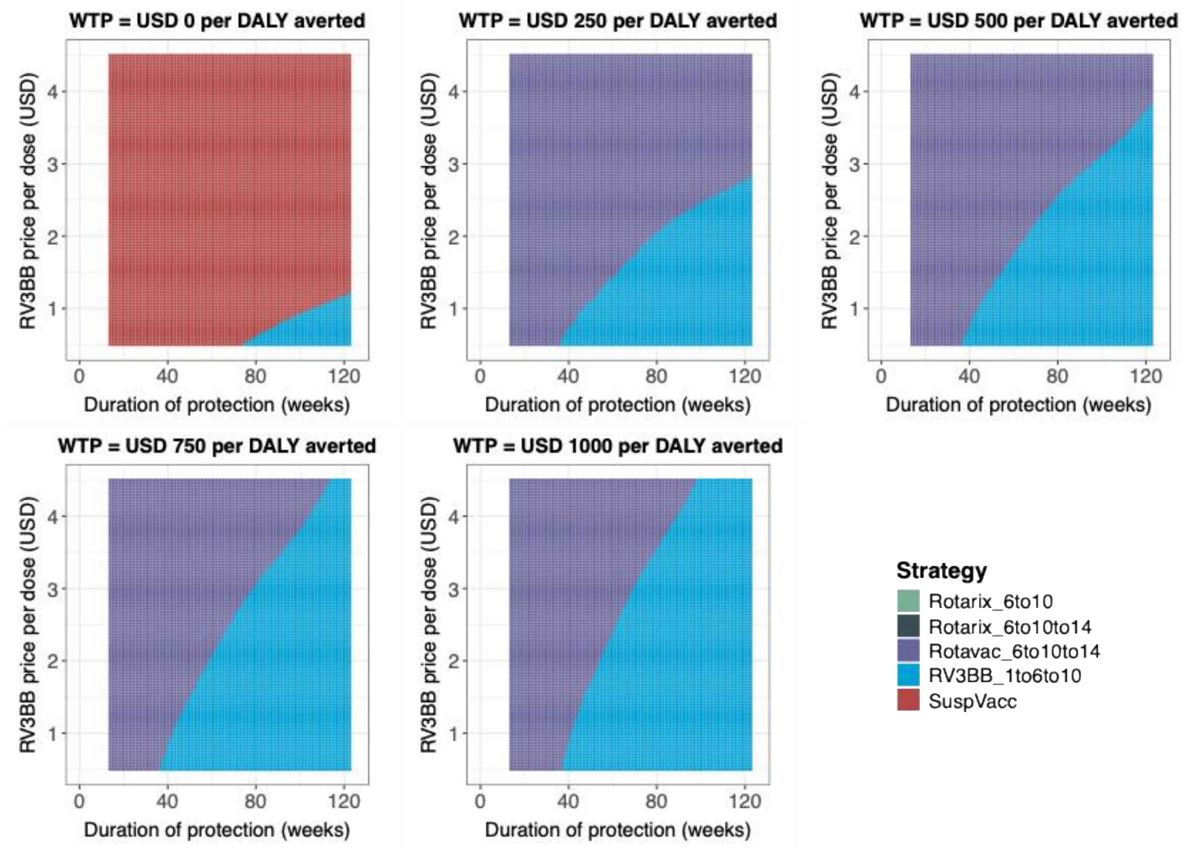

Figure S. ***15***). In the plots with WTP values above 0, the color patterns shifted slightly downward and to the right, indicating that a lower Rotavac price would require a correspondingly lower RV3-BB price for it to remain the preferred strategy at the same values.

In Ghana, assuming a lower Rotavac price per dose, Figure S. 16 and Figure S. 17 show that the color pattern marginally shifted compared with the base case.

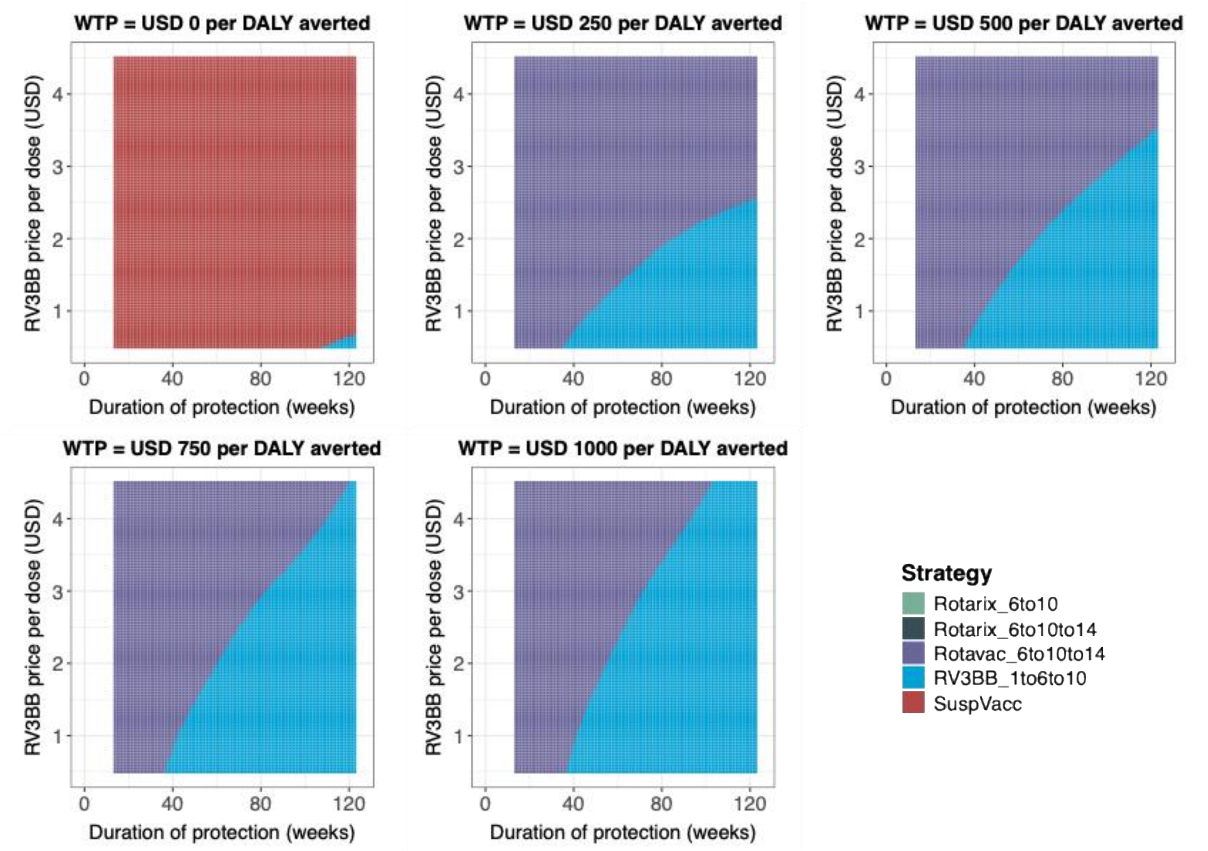

***Figure S. 14**: Two-way price and duration of protection probabilistic threshold analysis in Malawi from the government perspective assuming the price of Rotavac is $0.6 per dose with 20% wastage. Four strategies were compared with the current national immunization program and to each other under five willingness-to-pay values from $0 to $1000 per DALY averted. Y-axis presented price and x-axis presented duration of protection in weeks. Results are based on 5,000 simulations.*

Abbreviations: SuspVacc: Suspending vaccination, DALY: disability-adjusted life-year.

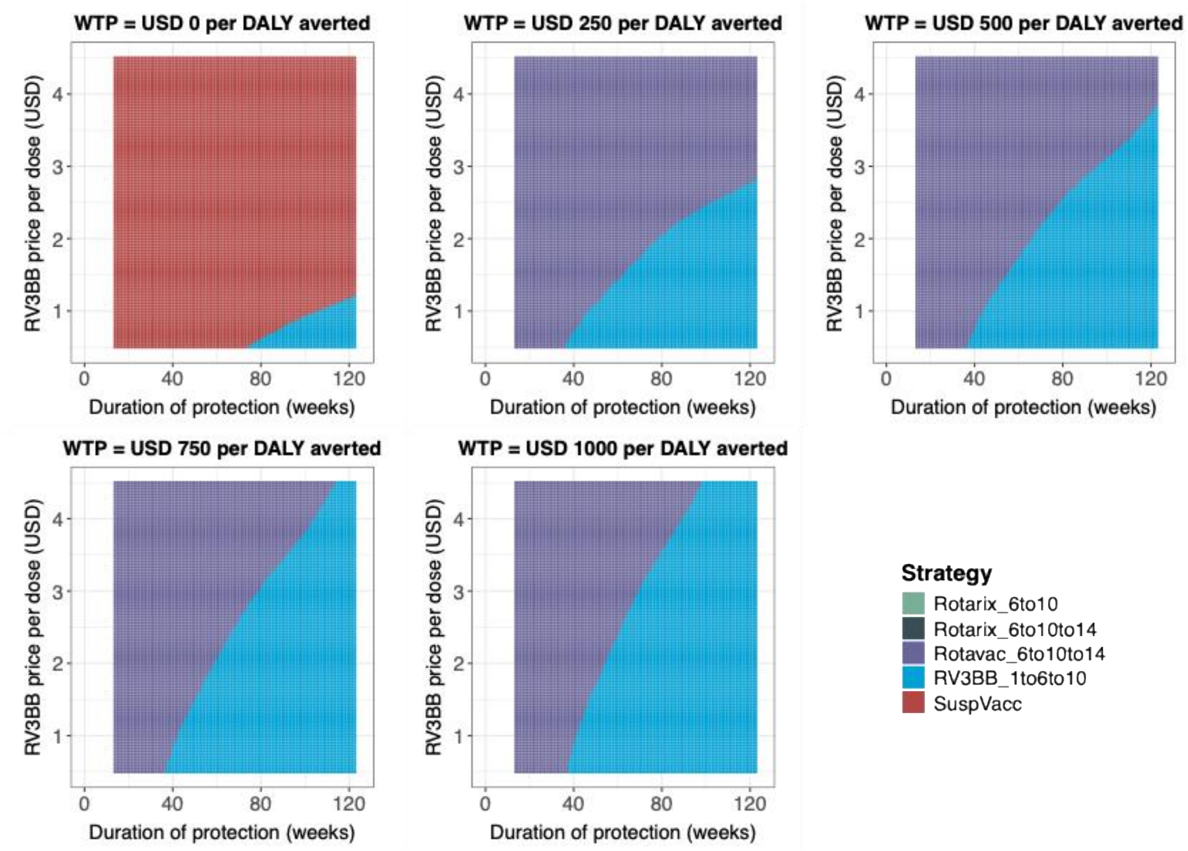

***Figure S. 15: Two-way price and duration of protection probabilistic threshold analysis in Malawi from a societal perspective assuming the price of Rotavac is $0.6 per dose with 20% wastage.*** *Four strategies were compared with the current national immunization program and to each other under five willingness-to-pay values from $0 to $1000 per DALY averted. Y-axis presented price and x-axis presented duration of protection in weeks. Results are based on 5,000 simulations.*

Abbreviations: SuspVacc: Suspending vaccination, DALY: disability-adjusted life-year.

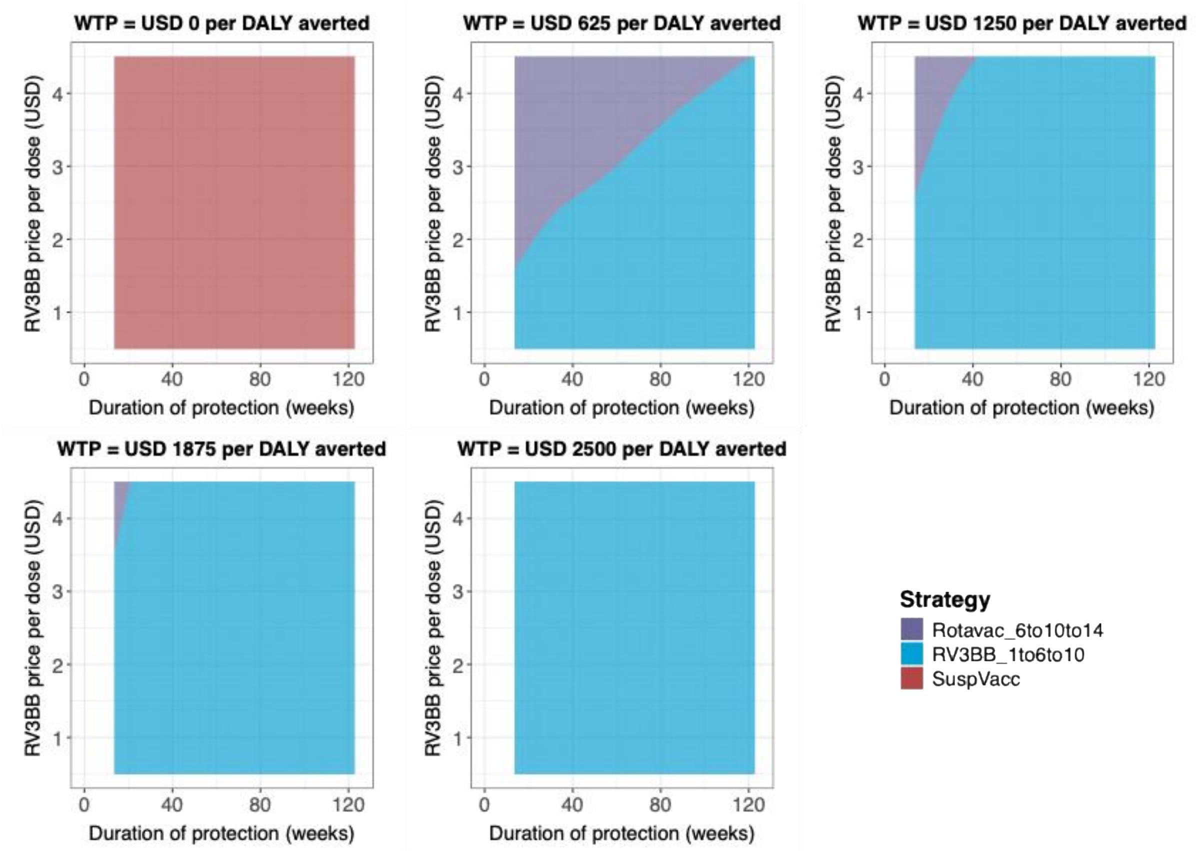

***Figure S. 16: Two-way price and duration of protection probabilistic threshold analysis in Ghana from the government perspective assuming the price of Rotavac is $0.6 per dose with 20% wastage****. Two strategies were compared with the current national immunization program and to each other under five willingness-to-pay values from $0 to $1000 per DALY averted. Y-axis presented price and x-axis presented duration of protection in weeks. Results are based on 5,000 simulations.*

Abbreviations: SuspVacc: Suspending vaccination, DALY: disability-adjusted life-year.

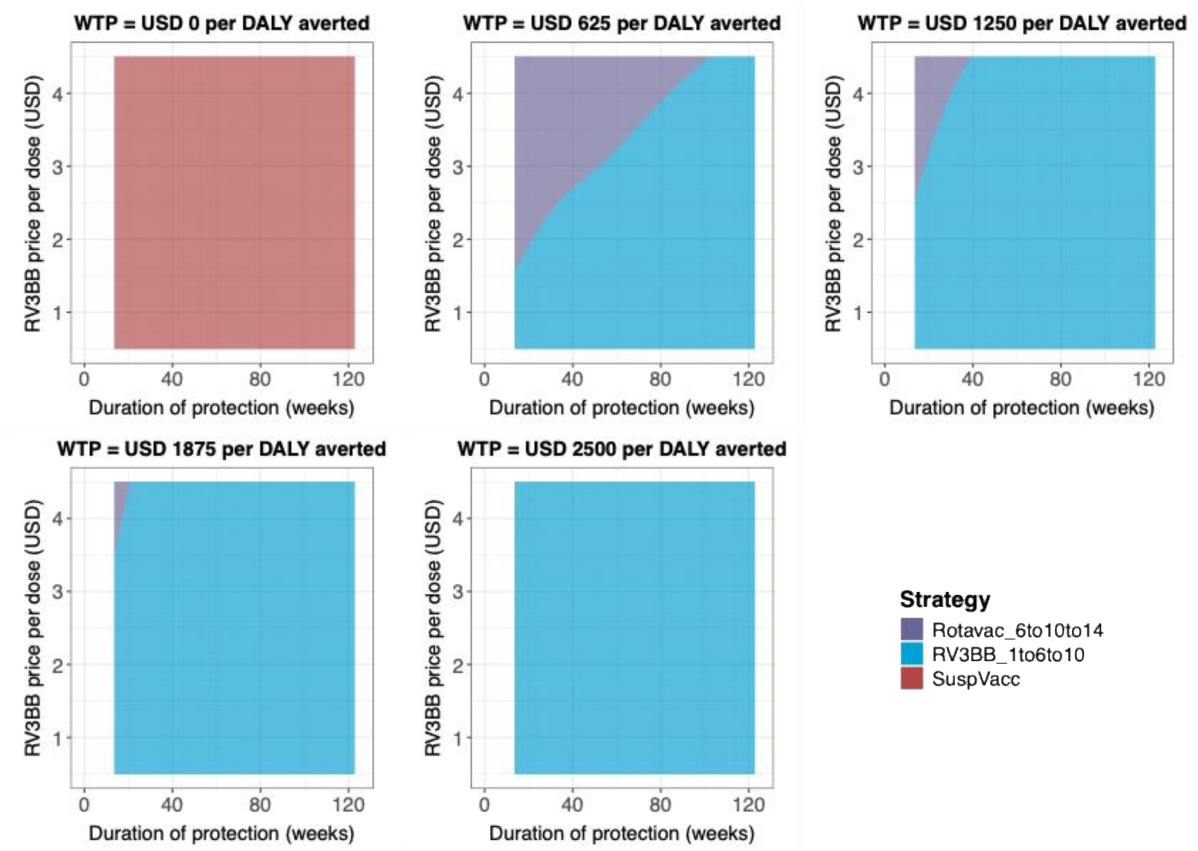

***Figure S. 17: Two-way price and duration of protection probabilistic threshold analysis in Ghana from a societal perspective assuming the price of Rotavac is $0.6 per dose with 20% wastage.*** *Two strategies were compared with the current national immunization program and to each other under five willingness-to-pay values from $0 to $1000 per DALY averted. Y-axis presented price and x-axis presented duration of protection in weeks. Results are based on 5,000 simulations.*

Abbreviations: SuspVacc: Suspending vaccination, DALY: disability-adjusted life-year.

##### 2.3.5 Threshold analysis for Malawi at initial self-financing phase, with copayment of $ 0.20 per dose for any rotavirus vaccine

We also conducted a sensitivity analysis for Malawi, reflecting its status in the Gavi initial self-financing phase. Under this scenario, Malawi pays a co-payment of $0.20 per dose for all vaccines.

The one-way sensitivity analysis (Figure S. 18) indicated that adding a third dose of Rotarix (dark green area) became the preferred strategy over Rotavac (purple area, Figure 2 in main text) when compared with the base case using full Gavi-negotiated prices. In the price sensitivity analysis (Figure S. 18, left plot), assuming RV3-BB was priced at $0.20 per dose, it can be the preferred strategy at values above $25 per DALY averted. If RV3-BB were not yet included in the Gavi procurement, it could still be preferred at prices ranging from $0.10 to $1.20 per dose across a range of WTP values. The baseline 2-dose Rotarix strategy also showed a limited chance of being preferred within a narrow range of WTP values (~$25–$40 per DALY averted) when the RV3-BB price was between $0.30 and $4.5 per dose (light green area).

When $0.20 per dose was assumed for all rotavirus vaccines, the 3-dose Rotarix strategy became the preferred option at WTP values above $75 per DALY averted and RV3-BB provided a duration of protection of less than 40 weeks (Figure S. 18, middle plot). The cost of switch had more impact compared to the base case analysis (Figure S. 18, right plot). The one-off switch cost was applied only when changing vaccine products, therefore, adding a third dose of Rotarix (dark green area) could become the preferred strategy when the switch cost ranged from $2.7 million to $5 million and the WTP values were between $75 and $400 per DALY averted. However, this assumption has limitations, as adding an additional Rotarix dose to current 2-dose Rotarix strategy may still incur some implementation costs, albeit likely lower than those associated with switching to a new vaccine product.

Two-way sensitivity analysis (Figure S. 19) consistently shows that when WTP values exceeded $0 per DALY averted, adding a third dose of Rotarix became the preferred strategy over Rotavac, in contrast to the base-case results. Nevertheless, RV3-BB remained the preferred strategy when priced low and providing longer protection (i.e. above 40 weeks), consistent with the base-case findings.

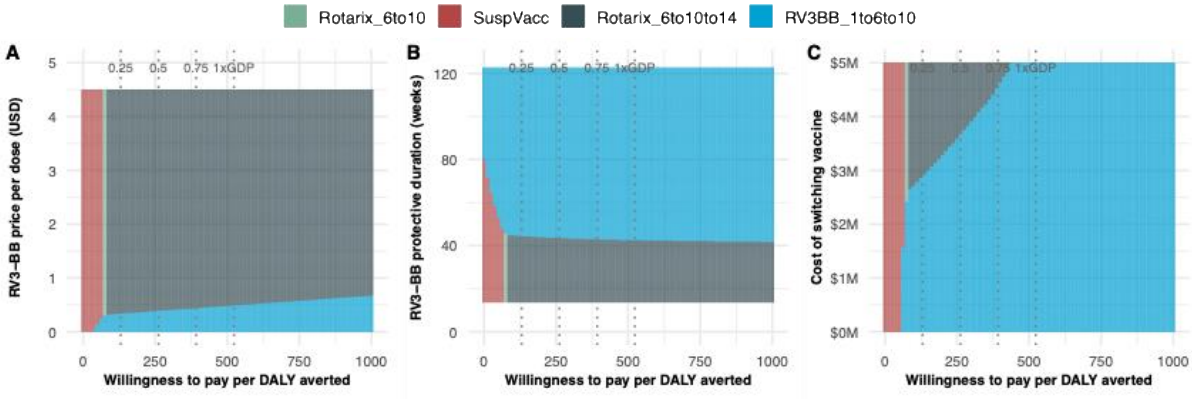

***Figure S. 18: One-way probabilistic threshold analysis in Malawi from the government perspective assuming Gavi co-financing****. The price of Rotarix and Rotavac per doses were fixed at $0.2. Except for the one-way probabilistic price threshold analysis, the price of RV-3BB was also fixed at $0.2 per dose. Four strategies were compared with the current national immunization program and to each other. Three parameters were evaluated independently: price (A), duration of protection (B) and switch cost (C). Results are based on 5,000 simulations.*

The 4 dotted vertical lines represented: 0.25, 0.5, 0.75 and 1 times Gross Domestic Product (GDP) per capita. Abbreviations: M: million, SuspVacc: Suspending vaccination.

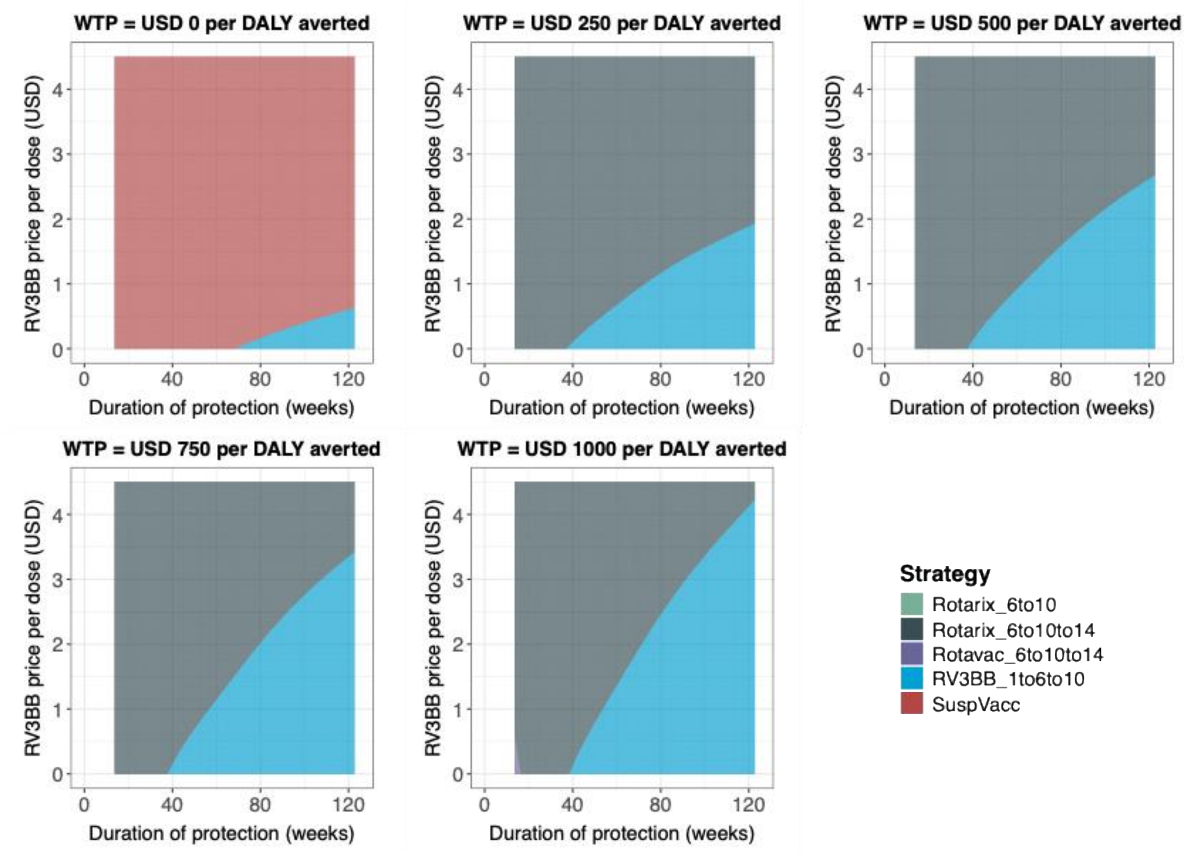

***Figure S. 19: Two-way price and duration of protection probabilistic threshold analysis in Malawi from the government perspective assuming Gavi co-financing**. Except RV3-BB, the prices of all vaccines were set as $0.2 per dose. Four strategies were compared with the current national immunization program and to each other under five willingness-to-pay values from $0 to $1000 per DALY averted. Y-axis presented price and x-axis presented duration of protection in weeks. Results are based on 5,000 simulations.*

Abbreviations: SuspVacc: Suspending vaccination, DALY: disability-adjusted life-year.

##### 2.3.6 Expected Value of Partially Perfect information (EVPPI)

The EVPPI was estimated for each uncertain input across varying WTP thresholds to identify key drivers of decision uncertainty. As shown in Figure S. 20, higher EVPPI values indicate greater influence of the corresponding parameter. We only presented the results from societal perspectives for both countries as indirect costs were also included in this analysis. Please note, in this analysis, the price of Rotarix, Rotavac and RV3-BB per doses were fixed at: $1.79, $0.7 and $0.7 respectively. The program switch cost was fixed at approximate $1million and the durability of RV3-BB were the same as base case.

In Malawi, EVPPI peaked at a value of $90 per DALY averted, indicating the greatest decision uncertainty at thresholds where the preferred strategy changes, consistent with results shown in Figure S. 20. Delivery cost was the most influential source of uncertainty, followed by hospitalization costs and outpatient costs for non-severe cases; the CFR of non–medically attended (non-MA) cases ranked fourth.

Similarly, in Ghana, delivery cost was also the top driver of uncertainty, while CFR for non-MA and MA cases were the second and third most influential parameters. Indirect outpatient costs for non-severe cases and assumptions regarding the outpatient-to-inpatient CFR ratio ranked fourth and fifth, respectively.

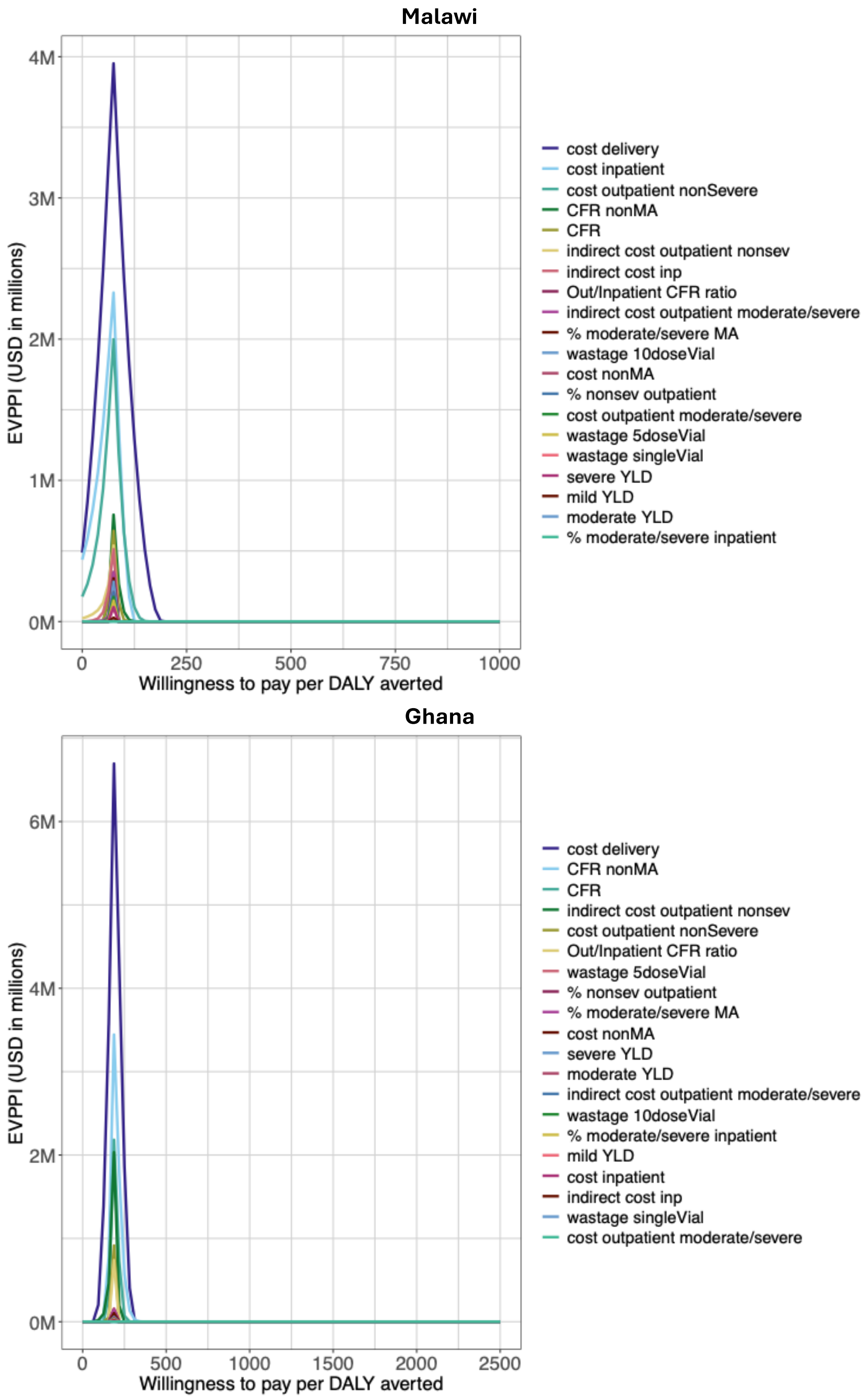

***Figure S. 20: Expected Value of Partially Prefect Information from a societal perspective in Malawi and Ghana.*** *Results are based on 5,000 simulations from a societal perspective.*

Abbreviation: DALY: disability-adjusted life-year, EVPPI: expected value of partially prefect information. YLD: Years Lived with Disability. CFR: case fatality ratio, MA: medically attended, prob: probability, YLD: years of life with disability

##### 2.3.7 Two-way threshold analysis on cost of delivery and price of RV3-BB

Aside from vaccine price and duration of protection, delivery cost per dose was identified as the most influential driver in both countries. Therefore, we conducted an additional two-way threshold analysis to examine how delivery cost per dose and vaccine price, together capturing the full variable cost of vaccination, affect the results.

In the base case, delivery costs were sampled uniformly from $0.88–$3.15 per dose (mean $2.02) in Malawi and $1.09–$3.65 per dose (mean $2.37) in Ghana. For this two-way threshold analysis, we explored a broader delivery cost range of $0–$4.50 per dose. From a societal perspective, the results are presented in Figure S. 21 for Malawi and Figure S. 22 for Ghana.

In Malawi, at a WTP value of $0 per DALY averted, RV3-BB was the preferred strategy when delivery costs were below $1 per dose and RV3-BB was priced below $1; otherwise, Rotavac was preferred (Figure S. 21, first plot). When delivery costs exceeded $1 per dose, suspension of vaccination became the preferred option. At a WTP value of $250 per DALY averted, RV3-BB was generally preferred at higher delivery costs ($3–$4 per dose) and a price of $1.10 per dose, while Rotavac was also preferred when delivery cost was $3 per dose. At WTP values above $250 per DALY averted, delivery costs had limited influence on the choice of preferred strategy, as both vaccines use 3-dose schedules.

In Ghana, at a WTP value of $0 per DALY averted, RV3-BB could be the preferred strategy when both its price and delivery costs were low (Figure S. 22). At a WTP value of $625 per DALY averted, RV3-BB was preferred when priced below $3.2 per dose with delivery costs up to $4.50 per dose. When delivery costs were below $4.4 per dose but the RV3-BB price exceeded $3.2 per dose, Rotavac became the preferred option. If delivery costs exceeded $4.4 per dose and RV3-BB was priced above $3.2 per dose, suspension of vaccination was preferred. At higher WTP values ($1,250–$2,500 per DALY averted), delivery costs had little impact on RV3-BB to be the preferred strategy.

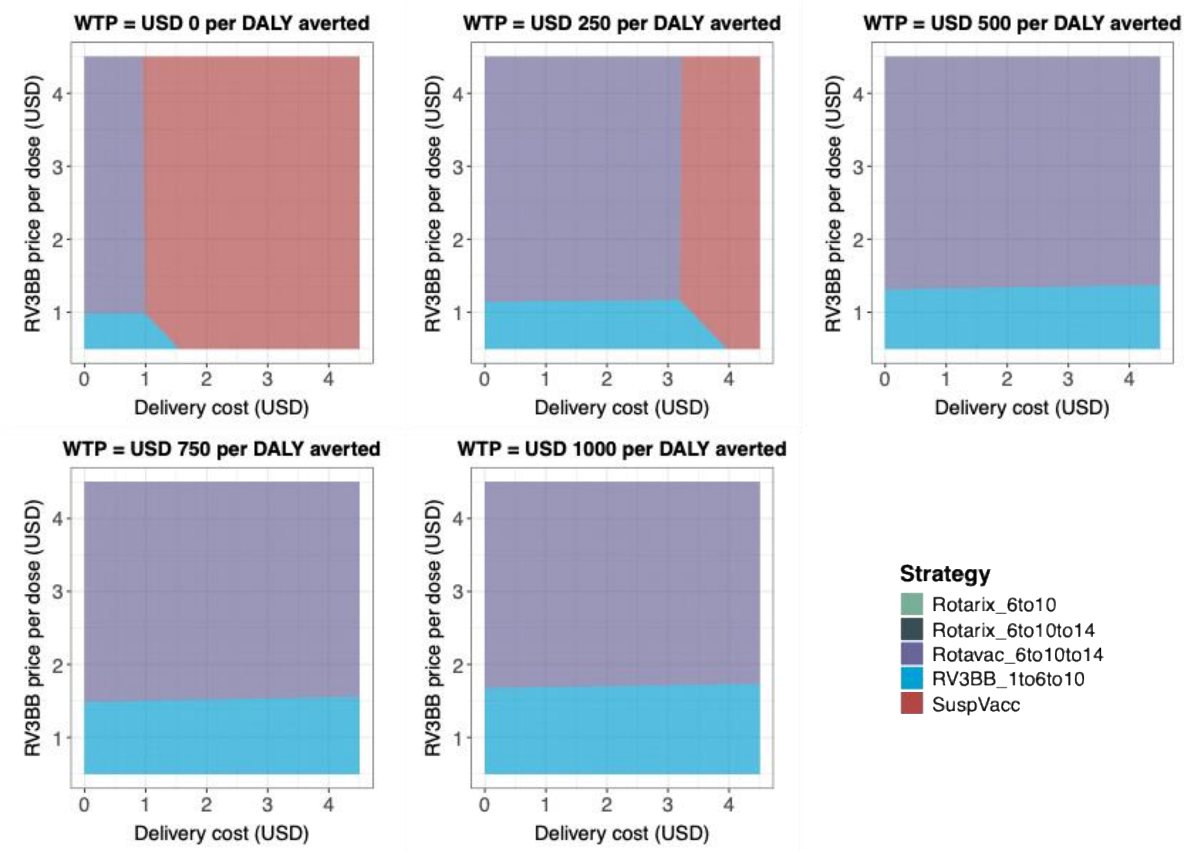

***Figure S. 21: Two-way price and delivery cost probabilistic threshold analysis in Malawi from a societal perspective****. Four strategies were compared with the current national immunization program and to each other under five willingness-to-pay values from $0 to $1000 per DALY averted. Y-axis presented price and x-axis presented duration of protection in weeks. Results are based on 5,000 simulations.*

Abbreviations: SuspVacc: Suspending vaccination, DALY: disability-adjusted life-year.

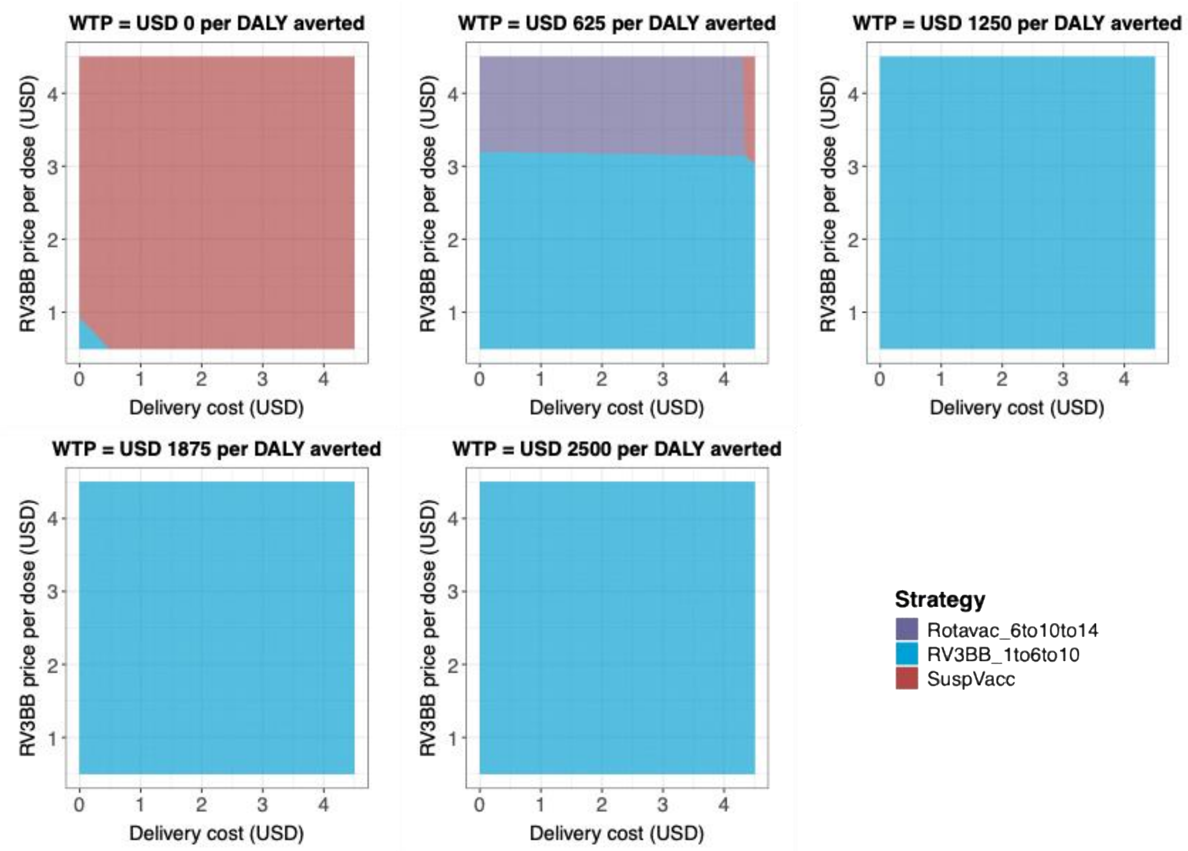

***Figure S. 22: Two-way price and delivery cost probabilistic threshold analysis in Ghana from a societal perspective****. Two strategies were compared with the current national immunization program and to each other under five willingness-to-pay values from $0 to $1000 per DALY averted. Y-axis presented price and x-axis presented duration of protection in weeks. Results are based on 5,000 simulations.*

Abbreviations: SuspVacc: Suspending vaccination, DALY: disability-adjusted life-year.
